# COVID-19 Surveillance in the Biobank at the Colorado Center for Personalized Medicine

**DOI:** 10.1101/2022.02.15.22271018

**Authors:** Randi K. Johnson, Katie M. Marker, David Mayer, Jonathan Shortt, David Kao, Kathleen C. Barnes, Jan T. Lowery, Christopher R. Gignoux

## Abstract

**Background:** Characterizing the experience and impact of the COVID-19 pandemic among various populations remains challenging due to the limitations inherent in common data sources such as the electronic health record (EHR) or convenience sample surveys.

**Objective:** To describe testing behaviors, symptoms, impact, vaccination status and case ascertainment during the COVID-19 pandemic using integrated data sources.

**Methods:** In summer 2020 and 2021, we surveyed participants enrolled in the Biobank at the Colorado Center for Personalized Medicine (CCPM, N = 180,599) about their experience with COVID-19. Prevalence of testing, symptoms, and the impacts of COVID-19 on employment, family life, and physical and mental health were calculated overall and by demographic categories. Using the Electronic Health Record (EHR), we compared COVID-19 case ascertainment and characteristics in the EHR versus the survey.

**Results:** Of the 25,063 survey respondents (13.9%), 42.5% had been tested for COVID-19 and of those, 12.8% tested positive. Nearly half of those tested had symptoms and/or had been exposed to someone who was infected. Young adults (18-29 years) and Hispanics were more likely to have positive tests compared to older adults and persons of other racial/ethnic groups. Mental health (54.6%) and family life (48.8%) were most negatively affected by the pandemic and more so among younger groups and women; negative impacts on employment were more commonly reported among Black respondents. After integration with EHR data up to the time of the survey completion, 4.0% of survey respondents (n=1,006) had discordant COVID-19 case status between the EHR and the survey. Using all longitudinal EHR and survey data, we identified 11,472 COVID-positive cases among Biobank participants (6.4%). In comparison to COVID-19 cases identified through the survey, EHR-identified cases were younger and more likely to be Hispanic.

**Conclusions:** Integrated data assets such as the Biobank at the CCPM are key resources for population health monitoring in response to public health emergencies, such as the COVID-19 pandemic.

## INTRODUCTION

The Coronavirus Disease 2019 (COVID-19) global pandemic has caused significant burden on the health and well-being of our families and communities. It has changed the way we work, socialize, and go about our daily lives. To date, over 888,000 Americans have died from COVID-19 and more than 49 million have been infected with the virus, many of whom have been hospitalized and/or suffered from a range of symptoms lasting from days to years [1]. Further, the burden of this disease, with respect to infection rates, hospitalizations, deaths and impacts on physical and mental health is not evenly distributed throughout the population. Understanding the nature and magnitude of this disease has been challenging due to the evolving nature of this virus, changing recommendations from public health around testing and self-quarantine, and our own health behaviors to avoid exposure.

As we strive to understand this novel virus in terms of risk and outcomes, it is important to assess the impact of COVID-19 among various populations including those who may experience serious versus mild effects from infection, those who experience symptoms but do not undergo testing, and those who never contract the disease. This broad inquiry requires multiple data sources. Electronic health records (EHR) are useful for capturing information on persons who seek medical care and/or become hospitalized due to COVID-19, and thus may reflect more severe cases [2-4]. However due to incomplete and unstructured data collection in EHRs, self-reported population surveys can provide information on persons with more mild disease who may opt not to seek medical care and those never infected [5]. Combining data sources from the EHR and surveys can mitigate limitations and biases inherent in each as well as optimize capture of the COVID-19 experience in a broader population.

We sought to characterize the experience and impact of the COVID-19 virus among a large and diverse group of persons enrolled in the Biobank at the Colorado Center for Personalized Medicine (CCPM), a collaborative initiative supported by UCHealth and the University of Colorado Anschutz Medical Campus. Specifically, we assessed the prevalence of testing and positive test results, the type and frequency of symptoms, health-care utilization, severity of disease, and the impacts of the pandemic on mental and physical health and employment. Uniquely, for this analysis, we were able to combine clinical data from the EHR, with self-reported information collected via an online survey that was offered to all Biobank participants.

We present here results from our analysis of self-reported survey data and clinical data recorded in the EHR for Biobank participants. By combining these unique data sources, we were able to capture more COVID-19+ cases and assess population differences in symptoms, healthcare utilization, severity (hospitalization), and personal impact. We also highlight the value of biobanks such as ours in facilitating rapid and comprehensive inquiries about emerging public health threats like COVID-19.

## METHODS

### Study Population

Enrollment in the CCPM Biobank is open to all UCHealth patients who are 18 years of age or older and able to provide consent for themselves through My Health Connection, the mobile EHR patient portal for UCHealth. Enrolled participants consent to use of their clinical data from the EHR and to being re-contacted about new research opportunities and to complete surveys. To date, the Biobank has enrolled over 200,000 adult participants from among the 2.5 million UCHealth patients across Colorado. Biobank participants are representative of the whole UCHealth population with respect to age, gender, race/ethnicity and co-morbidity status (**Multimedia Appendix 1**). For this study, all living Biobank participants with a valid email address were invited to complete an online survey about their experience with the COVID-19 pandemic. We linked survey responses with participants’ demographic and clinical data from the EHR that are captured and stored within Health Data Compass (Compass), the clinical data warehouse for UCHealth and the CCPM Biobank.

### Survey Development and Administration

We developed our survey based on an instrument developed by the International Common Disease Alliance (ICDA) [6] early in the pandemic. Our survey included questions about testing for COVID-19, test results, symptoms related to COVID-19 infection, health care utilization following a positive test and/or symptoms, underlying health conditions, the impact of COVID-19 on health and well-being, potential household exposure to COVID-19, and current smoking behaviors (**Multimedia Appendix 2**). We created the survey in REDCap [7], a HIPAA-compliant database and research management platform, and created unique survey links for each Biobank participant. Personal invitations to complete the survey were sent by email to all participants beginning in June 2020 with a follow-up reminder to non-responders within 2 weeks. We repeated the process in October 2020 to all newly enrolled participants between June and October 2020. We revised the survey in March 2021 to include additional questions on vaccine uptake, adverse reactions to the vaccine and long-term symptoms post infection (**Multimedia Appendix 3**). The revised survey was sent to all participants who hadn’t responded to the initial survey and newly enrolled participants through May 2021. In total, survey invites were sent to 180,599 individual participants over the course of 15 months.

### COVID-19 Case and Severity Definitions: Survey and EHR

Survey respondents who reported receiving a positive COVID-19 test result were considered a “confirmed case” of COVID-19. Self-reported cases also reported whether the respondent tested positive for COVID-19, saw a doctor in-person or through telehealth, visited the emergency room, were hospitalized overnight, stayed home/isolated, or did nothing different. We looked at severity either in terms of hospitalization due to COVID-19 or death after COVID-19. Respondents who reported having one or more overnight stays in the hospital were considered to be ‘hospitalized’.

Positive cases were identified in the EHR using ICD-10 diagnosis codes, healthcare encounter types, and encounter primary diagnoses. Participants who received an ICD-10 diagnosis code of U07.1 or at least one of 11 COVID-19 specific encounter primary diagnoses (**Multimedia Appendix 4**) were considered an “EHR-confirmed case”. Participants who were hospitalized in a UCHealth hospital overnight during the 3 days before or up to 21 days after their COVID-19 diagnosis date and who had at least one of 64 COVID-19 related encounter primary diagnoses (**Multimedia Appendix 5**) were considered to be “EHR-hospitalized.” To compare positive cases identified from the EHR and survey, we examined the number of hospitalized cases that were discordant between these data sources.

All-cause mortality data stored in the Health Data Compass clinical data warehouse include the cause of death as certified by a physician or coroner/medical examiner, related ICD-10 cause of death codes generated by Centers for Disease Control, and age at death. These data are obtained through routine linkage of UCHealth patients with the vital statistics/death certificates provided by the Department of Vital Statistics at the Colorado Department of Public Health and the Environment (CDPHE). Accounting for the ∼3-month lag time to register certificates, map ICD-10 cause of death codes, and update the clinical databases, the ascertainment of mortality among UCHealth patients for this analysis is nearly 95% complete.

### Other Definitions

Age and race-ethnicity were determined from the EHR.. Race and ethnic indicators were extracted as encoded in the EHR and categorized into 4 racial-ethnic groups to preserve >10 individuals in each group in all analyses, including: non-Hispanic White, non-Hispanic Black, any Hispanic, other.

### Statistical Analysis

We generated descriptive statistics to characterize our study population and responses to survey questions. We also stratified respondents with respect to COVID-19 infection status based on reported test status and symptomology. We compared COVID-19+ individuals that were identified via the survey and via the EHR by demographics and severity (overnight hospitalization and death). We investigated case status and hospitalization misclassification in both the survey and EHR by comparing those who were discordant in the survey and EHR. We calculated differences between groups using chi-square and t-test statistics for categorical and continuous measures, respectively. As expected, due to very large sample size in the study, most comparisons were statistically significant at a two-sided alpha < 0.05. Therefore, we focus results and interpretation on effect sizes and corresponding standard error of the estimate.

## RESULTS

### Survey response

Of 180,599 biobank participants with valid email addresses, 25,063 completed at least one survey and had complete demographic information (response rate = 13.9%, **Figure 1**). Compared to non-respondents (**Multimedia Appendix 6**), respondents were older (mean age = 55.0 years vs. 48.6 years, *P*<.001) and enriched for a higher proportion of females (62.6% vs. 59.0%, *P*<.001) and individuals of non-Hispanic White race-ethnicity (87.4% vs. 77.1%, *P*<.001).

**Figure 1:**
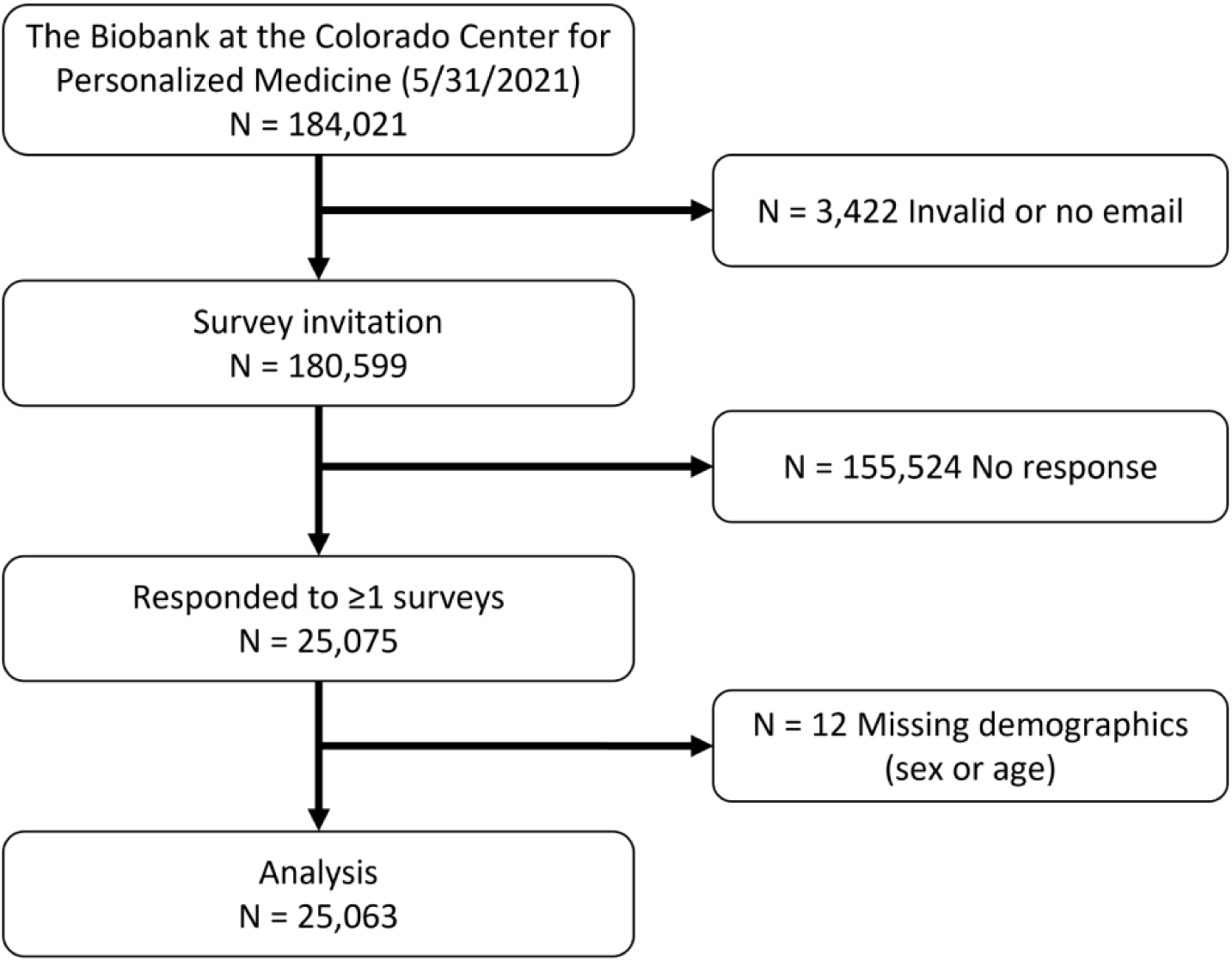
The Biobank at the Colorado Center for Personalized Medicine COVID-19 Survey Population.

### COVID-19 Testing

Among all survey respondents, 42.5% (n=10,661) reported being tested for COVID-19. The most common reasons for testing were having symptoms (29.5%), exposure to someone who tested positive for COVID-19 (18.5%), doctor recommendation (14.7%), requirement of employer (8.9%), and recent international travel (3.4%). An additional 40.8% of individuals tested, reported ‘other’ reasons for testing that included having surgery or other medical procedure, planned travel, desire or need to be around large groups or family members and work-site offerings for testing.

Of those tested, 1366 (12.8%) tested positive for COVID-19 (**Table 1**) and were considered confirmed cases. The distributions of age, sex, race-ethnicity, college education, number of symptoms, number of pre-existing comorbidities, overall health status, and exposure to a household member who tested positive for COVID-19 were different across the three groups of those who tested positive, tested negative, and were not tested (all *P*<.001). Young adults (ages 18-29 years) were overrepresented among the tested-positive group, representing 10.7% of those who tested positive compared to 6.7% of those who tested negative and 5.1% of those who were not tested (*P* for trend <.001). Similarly, individuals of Hispanic race-ethnicity were overrepresented in the tested-positive group at 9.2%, compared to 5.7% of those who tested negative and 4.3% of those who were not tested. Individuals who tested positive were also more likely to report symptoms, household exposure to COVID-19 and poor health status (Table 1, all *P*<.001).

**Table 1:**
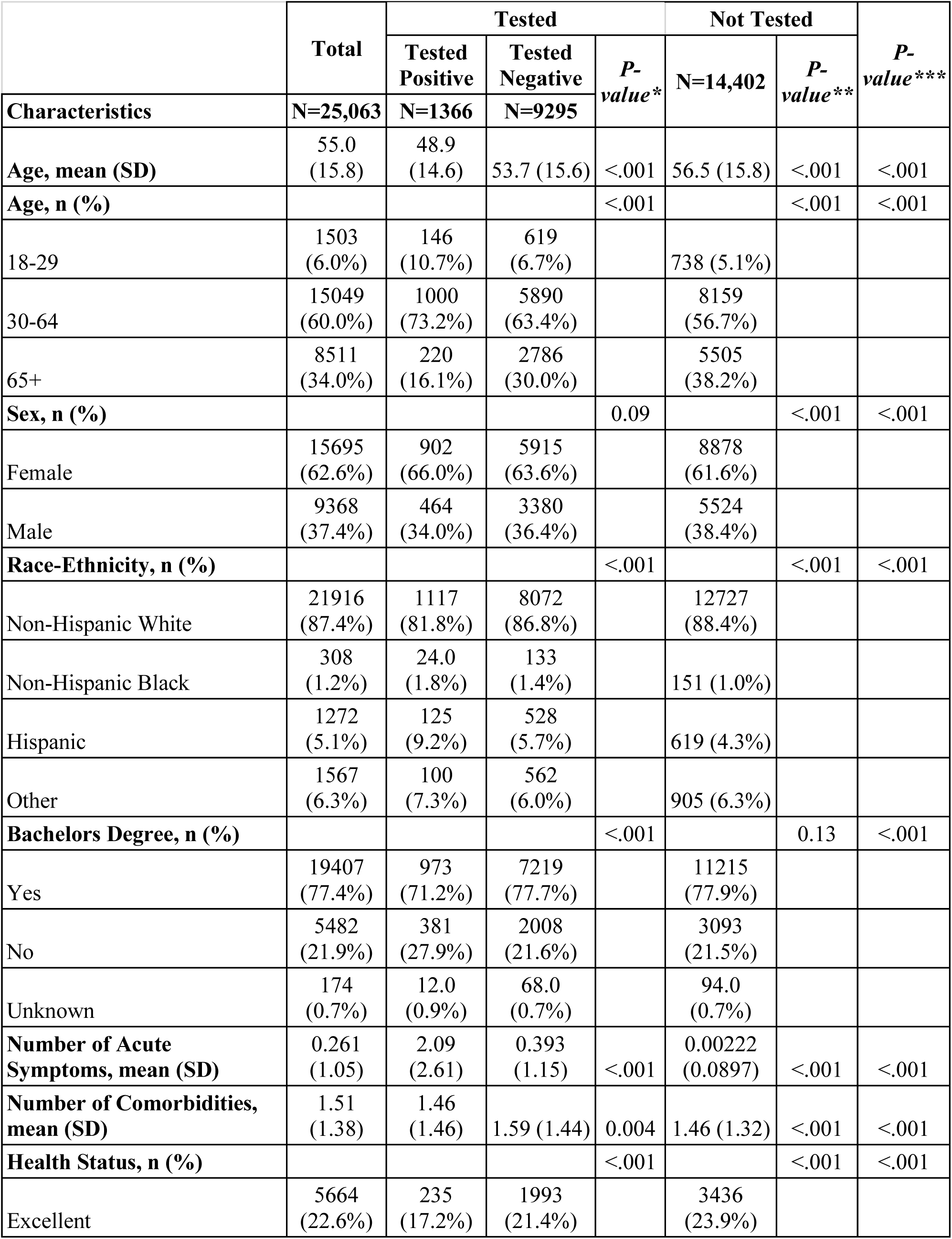
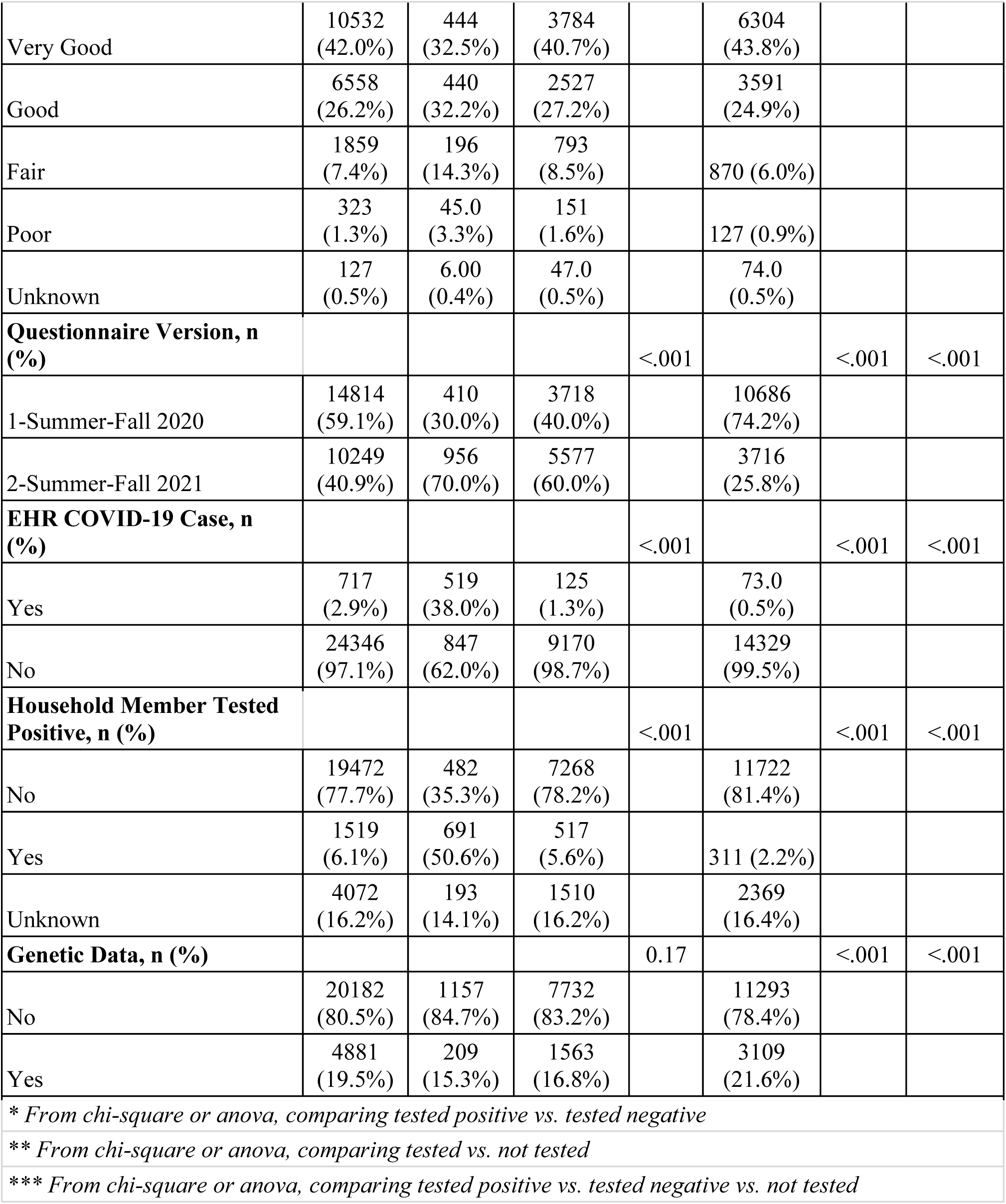
COVID-19 Testing in the Biobank among Survey Respondents.

|  | Total | Tested |  |  | Not Tested |  | *P-value\*\*\** |
| --- | --- | --- | --- | --- | --- | --- | --- |
|  |  | Tested Positive | Tested Negative | *P-value\** | N=14,402 | *P-value\*\** |  |
| **Characteristics** | **N=25,063** | **N=1366** | **N=9295** |  |  |  |  |
| **Age, mean (SD)** | 55.0<br>(15.8) | 48.9<br>(14.6) | 53.7 (15.6) | <.001 | 56.5 (15.8) | <.001 | <.001 |
| **Age, n (%)** |  |  |  | <.001 |  | <.001 | <.001 |
| 18-29 | 1503<br>(6.0%) | 146<br>(10.7%) | 619<br>(6.7%) |  | 738 (5.1%) |  |  |
| 30-64 | 15049<br>(60.0%) | 1000<br>(73.2%) | 5890<br>(63.4%) |  | 8159<br>(56.7%) |  |  |
| 65+ | 8511<br>(34.0%) | 220<br>(16.1%) | 2786<br>(30.0%) |  | 5505<br>(38.2%) |  |  |
| **Sex, n (%)** |  |  |  | 0.09 |  | <.001 | <.001 |
| Female | 15695<br>(62.6%) | 902<br>(66.0%) | 5915<br>(63.6%) |  | 8878<br>(61.6%) |  |  |
| Male | 9368<br>(37.4%) | 464<br>(34.0%) | 3380<br>(36.4%) |  | 5524<br>(38.4%) |  |  |
| **Race-Ethnicity, n (%)** |  |  |  | <.001 |  | <.001 | <.001 |
| Non-Hispanic White | 21916<br>(87.4%) | 1117<br>(81.8%) | 8072<br>(86.8%) |  | 12727<br>(88.4%) |  |  |
| Non-Hispanic Black | 308<br>(1.2%) | 24.0<br>(1.8%) | 133<br>(1.4%) |  | 151 (1.0%) |  |  |
| Hispanic | 1272<br>(5.1%) | 125<br>(9.2%) | 528<br>(5.7%) |  | 619 (4.3%) |  |  |
| Other | 1567<br>(6.3%) | 100<br>(7.3%) | 562<br>(6.0%) |  | 905 (6.3%) |  |  |
| **Bachelors Degree, n (%)** |  |  |  | <.001 |  | 0.13 | <.001 |
| Yes | 19407<br>(77.4%) | 973<br>(71.2%) | 7219<br>(77.7%) |  | 11215<br>(77.9%) |  |  |
| No | 5482<br>(21.9%) | 381<br>(27.9%) | 2008<br>(21.6%) |  | 3093<br>(21.5%) |  |  |
| Unknown | 174<br>(0.7%) | 12.0<br>(0.9%) | 68.0<br>(0.7%) |  | 94.0<br>(0.7%) |  |  |
| **Number of Acute Symptoms, mean (SD)** | 0.261<br>(1.05) | 2.09<br>(2.61) | 0.393<br>(1.15) | <.001 | 0.00222<br>(0.0897) | <.001 | <.001 |
| **Number of Comorbidities, mean (SD)** | 1.51<br>(1.38) | 1.46<br>(1.46) | 1.59 (1.44) | 0.004 | 1.46 (1.32) | <.001 | <.001 |
| **Health Status, n (%)** |  |  |  | <.001 |  | <.001 | <.001 |
| Excellent | 5664<br>(22.6%) | 235<br>(17.2%) | 1993<br>(21.4%) |  | 3436<br>(23.9%) |  |  |
| Very Good | 10532<br>(42.0%) | 444<br>(32.5%) | 3784<br>(40.7%) |  | 6304<br>(43.8%) |  |  |
| Good | 6558<br>(26.2%) | 440<br>(32.2%) | 2527<br>(27.2%) |  | 3591<br>(24.9%) |  |  |
| Fair | 1859<br>(7.4%) | 196<br>(14.3%) | 793<br>(8.5%) |  | 870 (6.0%) |  |  |
| Poor | 323<br>(1.3%) | 45.0<br>(3.3%) | 151<br>(1.6%) |  | 127 (0.9%) |  |  |
| Unknown | 127<br>(0.5%) | 6.00<br>(0.4%) | 47.0<br>(0.5%) |  | 74.0<br>(0.5%) |  |  |
| **Questionnaire Version, n (%)** |  |  |  | <.001 |  | <.001 | <.001 |
| 1-Summer-Fall 2020 | 14814<br>(59.1%) | 410<br>(30.0%) | 3718<br>(40.0%) |  | 10686<br>(74.2%) |  |  |
| 2-Summer-Fall 2021 | 10249<br>(40.9%) | 956<br>(70.0%) | 5577<br>(60.0%) |  | 3716<br>(25.8%) |  |  |
| **EHR COVID-19 Case, n (%)** |  |  |  | <.001 |  | <.001 | <.001 |
| Yes | 717<br>(2.9%) | 519<br>(38.0%) | 125<br>(1.3%) |  | 73.0<br>(0.5%) |  |  |
| No | 24346<br>(97.1%) | 847<br>(62.0%) | 9170<br>(98.7%) |  | 14329<br>(99.5%) |  |  |
| **Household Member Tested Positive, n (%)** |  |  |  | <.001 |  | <.001 | <.001 |
| No | 19472<br>(77.7%) | 482<br>(35.3%) | 7268<br>(78.2%) |  | 11722<br>(81.4%) |  |  |
| Yes | 1519<br>(6.1%) | 691<br>(50.6%) | 517<br>(5.6%) |  | 311 (2.2%) |  |  |
| Unknown | 4072<br>(16.2%) | 193<br>(14.1%) | 1510<br>(16.2%) |  | 2369<br>(16.4%) |  |  |
| **Genetic Data, n (%)** |  |  |  | 0.17 |  | <.001 | <.001 |
| No | 20182<br>(80.5%) | 1157<br>(84.7%) | 7732<br>(83.2%) |  | 11293<br>(78.4%) |  |  |
| Yes | 4881<br>(19.5%) | 209<br>(15.3%) | 1563<br>(16.8%) |  | 3109<br>(21.6%) |  |  |
| *\* From chi-square or anova, comparing tested positive vs. tested negative* |  |  |  |  |  |  |  |
| *\*\* From chi-square or anova, comparing tested vs. not tested* |  |  |  |  |  |  |  |
| *\*\*\* From chi-square or anova, comparing tested positive vs. tested negative vs. not tested* |  |  |  |  |  |  |  |

### COVID-19 Case Symptomology

Out of the 1,366 COVID-19+ individuals identified from the survey, 1,154 (84.4%) individuals had at least one of the following COVID-19 related symptoms since February 2020: cough, fever over 99.9°F, general tiredness/fatigue, muscle/body aches, runny nose, difficulty breathing/shortness of breath, loss of sense of smell or taste, stomach or gastrointestinal (GI) problems (**Figure 2**). However, only 48.4% reported at least one symptom 14 days before or after a positive COVID-19 test (n = 661). The number of symptoms individuals reported is relatively even from one to eight symptoms, ranging from 3.7% (n = 50) reporting all eight symptoms and 8.4% (n = 115) reporting four symptoms (Figure 2A). General tiredness/fatigue and muscle/body aches were the most commonly reported symptoms within 14 days of a positive COVID-19 test, at 37.8% and 31.2%, respectively (Figure 2B). Next most common was loss of sense of smell or taste, with 397 (29.1%) reporting within 14 days of a positive COVID-19 test (Figure 2B). However, an additional 283 individuals reported this symptom outside the 28-day window. A quarter of individuals reported a cough within 14 days of a positive COVID-19 test (n = 346) and 22.7% and 22.1% reported difficulty breathing/shortness of breath and a runny nose, respectively (Figure 2B). Only 17.1% of individuals (n = 234) reported stomach or GI problems (Figure 2B). The remainder (51.6%) reported no symptoms within 14 days before or after their COVID-19 positive test (n = 705). There were no significant differences in asymptomatic cases compared to symptomatic cases (having at least one symptom) when comparing by age, sex, or race/ethnicity (Figure 2C-E).

**Figure 2:**
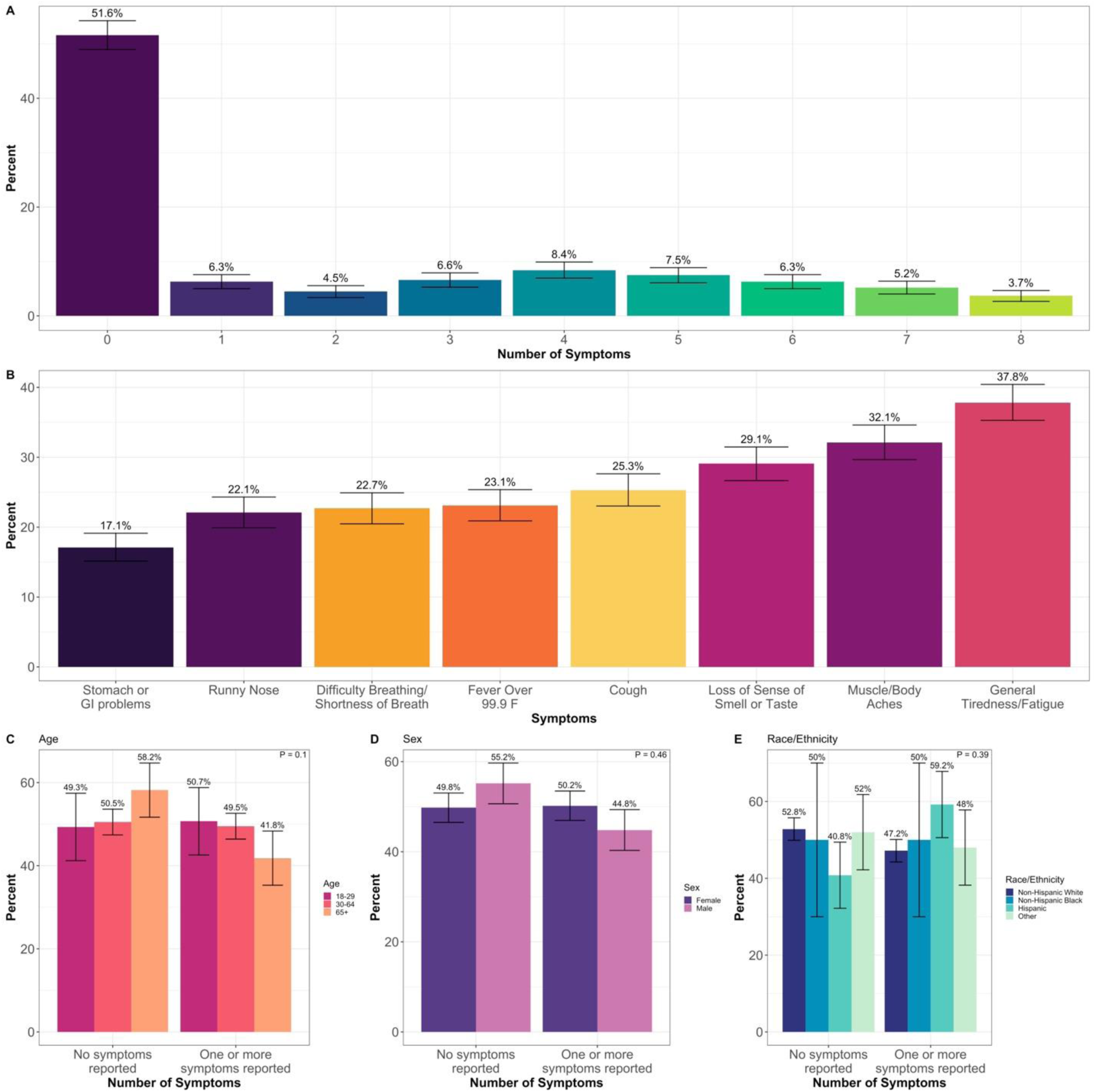
Symptomology among COVID-19 cases. Each symptom was reported 14 days before or 14 days after a positive COVID-19 test. A) Number of symptoms reported among COVID-19 cases. B) Percent of COVID-19 positive cases that reported each symptom. Comparing asymptomatic cases compared to symptomatic cases (at least one symptom) by C) age, D) sex, E) race/ethnicity. P = *p*-value from Pearson’s chi-square test for different distributions across demographic groups. Error bars indicate the 95% confidence interval for the percent point estimate.

### Health Behaviors and Impact on Healthcare System

To assess health behaviors among COVID-19 cases and the potential impact on the healthcare system, we asked these individuals what they did as a result of testing positive (Figure 3A). Eighty-one percent of the 1,366 respondents with positive tests stayed home and self-isolated (n = 1108), and 5.6% did not report any changes in behavior (n = 76) (Figure 3A). Of those who did not change behavior, 82.9% did not have any symptoms reported 14 days before or after their COVID-19 test (n = 63). Among the 1,366 positives, 625 (45.8%) of those sought out at least one form of medical care. One hundred ninety (14%) saw a doctor at an in-person visit, 454 (33.2%) saw a doctor via telehealth, 14.2% went to the emergency room (n = 194) and 7.9% had an overnight stay in a hospital (n = 108) (Figure 3A). A subset of 229 (16.7%) individuals reported being tested at a UCHealth facility (n = 229, 16.7%) vs. 213 (15.6%) outside UCHealth with no response from 924 respondents. Of the 229 respondents who said they tested positive at a UCHealth facility, only 59.8% (n = 137), were identified as a ‘case’ within the EHR. There is a high rate of missingness for the question on who performed the test (67.6%), so there may be confusion by participants about who supplied the COVID-19 test.

**Figure 3:**
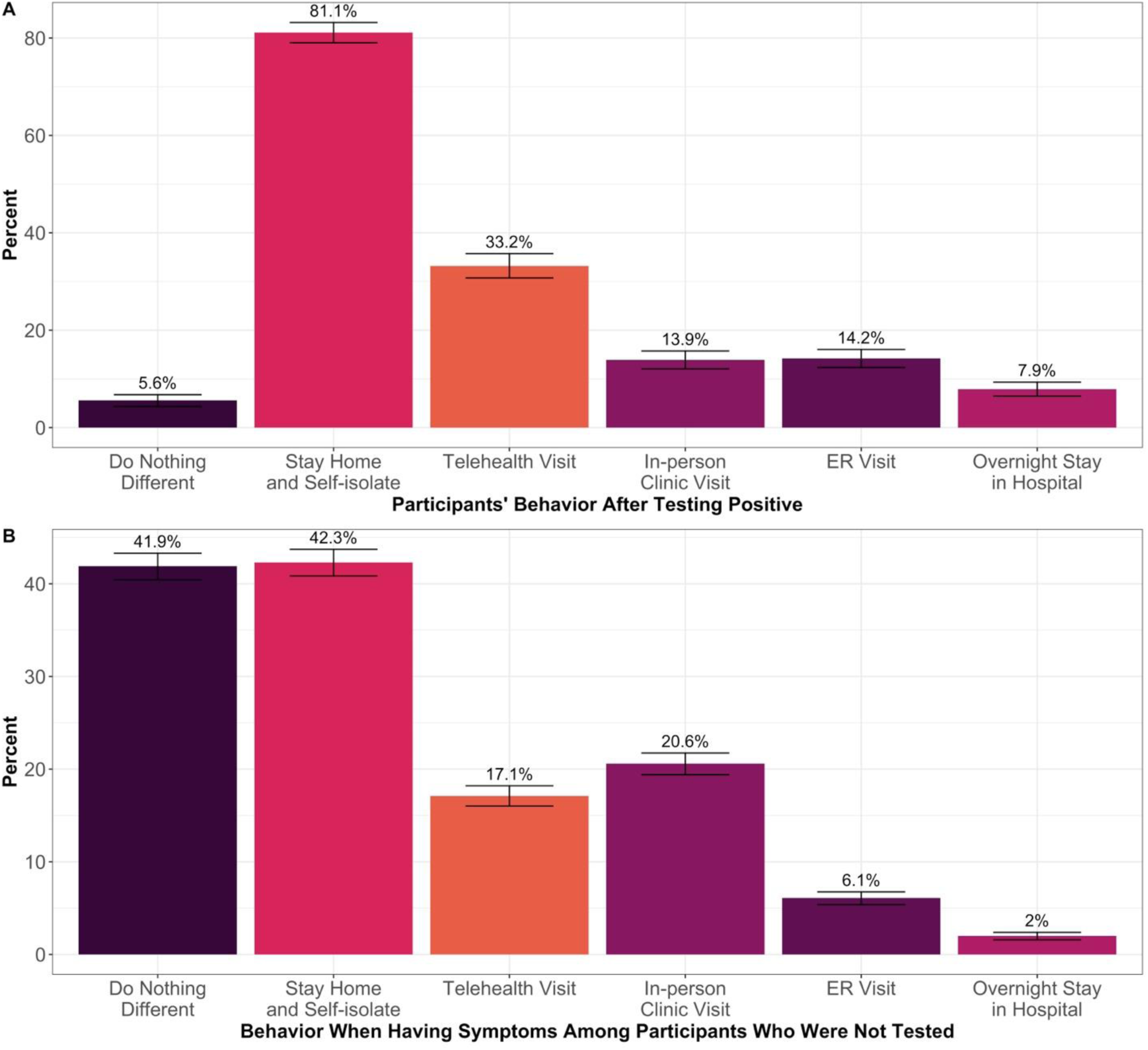
The impact of COVID-19 on the health care system. A) Participants’ behavior after testing positive for COVID-19, B) Participants’ behavior when having symptoms, among those with at least one symptom whom did not get tested for COVID-19. Error bars indicate the 95% confidence interval for the percent point estimate.

Among respondents who were not tested but reported having at least 1 COVID-related symptom, 1901 (41.9%) said they did nothing different, whereas 1920 (42.3%) stayed home and self-isolated (Figure 3B). A third (n = 1,515) sought out at least one form of medical care, 934 (20.6%) had an in-person clinic visit, 77 (17.1%) had a telehealth clinic visit, 275 (6.1%) went to the ER, and 90 (2.0%) had an overnight stay in the hospital (Figure 3B).

### Impact of the COVID-19 Pandemic

The impact of the COVID-19 pandemic on employment, family life, mental health or physical health was largely negative, with more than 75% of respondents (n=18,861) reporting a negative impact from the COVID-19 pandemic in at least one of these domains, compared to 23% of respondents (n=5,856) reporting a positive impact in at least one domain (*P*<.001). Mental health and family life were most negatively affected by the pandemic, at 54.6% (n=13,688) and 48.8% (n=12,233) of respondents reporting a negative impact, respectively. Negative impact in other two domains was lower at 28.2% (n=7,059) for physical health and 21.2% (n=5,320) for employment (*P*<.001).

The impact of the COVID-19 pandemic was not equal across groups by age, race-ethnicity, sex, and COVID-19 testing status (max *P*=.006, **Figure 4**). A higher proportion of young adults reported a negative mental health impact (74.9%, 95%CI = 72.7-77.1%) than adults aged 30-64 years (60.7%, 95%CI = 59.6-61.5%) and older adults, 65+ years (41.1%, 95%CI = 40.1-42.2%). A similar linear trend across age groups was seen for the negative impact of the pandemic on employment and physical health (Figure 4A). Using self-reported race-ethnicity as captured in the EHR, a higher proportion of non-Hispanic Black respondents reported a negative impact on their employment (35.8%, 95%CI = 30.4-41.2%) compared to other race-ethnic groups (all proportions < 26.1%, Figure 2B). Women reported greater negative impact in all four domains compared to men, at (respectively) 23.4% versus 18.3% in employment, 50.5% versus 47.0% in family life, 60.3% versus 46.0% in mental health, and 30.7% versus 24.4% in physical health (Figure 4C, all *P*<.001). Respondents who tested positive for COVID-19 reported a higher negative impact on their physical health (55.0%, 95%CI = 52.3-57.6%) than those who tested negative (30.9%, 95%CI = 30.0-31.9%) and those who did not report a COVID-19 test (24.2%, 95%CI = 23.5-24.9%, Figure 4D).

**Figure 4:**
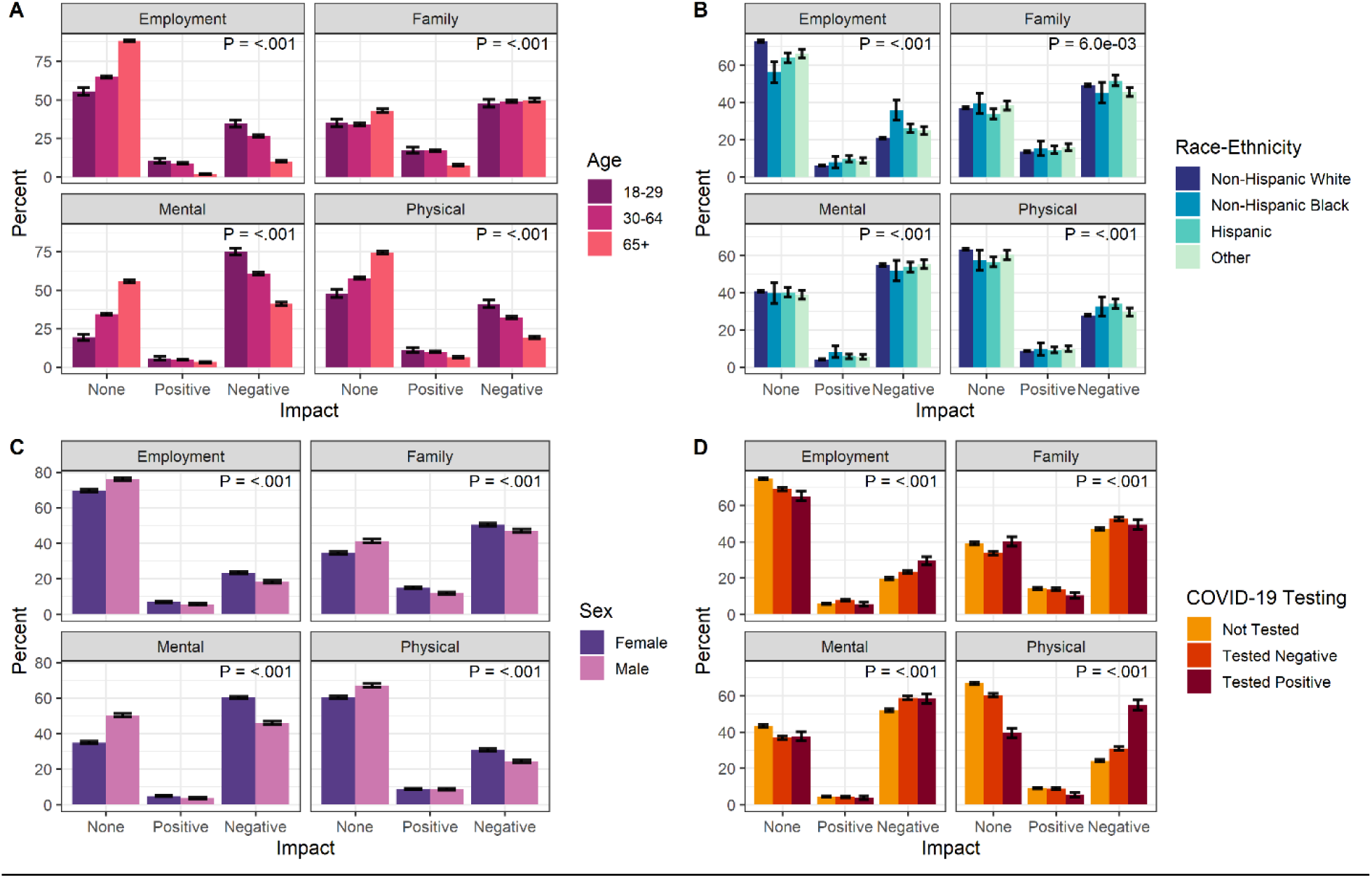
The impact of COVID-19 on employment, family life, mental and physical health by A) age, B) race-ethnicity, C) sex, and D) COVID-19 test status. P = *p*-value from Pearson’s chi-square test for different distributions across impact and demographic groups. Error bars indicate the 95% confidence interval for the percent point estimate.

### COVID-19 Vaccination

In our second round of the survey (administered Spring/Summer 2021), we added questions about COVID-19 vaccination. Of the 10,249 individuals who responded, 95.3% had received the vaccine (n = 9,770). Younger people were less likely to have received a vaccine: 7.5% of those aged 18-29 years did *not* receive a vaccine compared to 4.9% of individuals ages 30-64 years and 2.0% of individuals 65+ (*P*<.001, **Figure 5A**). Women were slightly less likely to receive a vaccine (4.5% of women vs. 3.3% of men; *P* = 0.003, **Figure 5B**). Vaccination rate was very similar across race/ethnicity categories, with 4.1% of non-Hispanic Whites, 4.3% of non-Hispanic Blacks, 4.1% of Hispanics, and 3.3% of those in the other race category not receiving vaccines (*P* = 0.8, Figure 5C). The median income of the home 3-digit zip code was lower for unvaccinated participants: $67,800 in the unvaccinated compared to $71,500 in the vaccinated (*P*<.001). The median percent of the population who received a bachelor’s degree by 3-digit zip code was lower for unvaccinated participants (37.7%) compared to vaccinated participants (47.3%), *P*<.001.

**Figure 5:**
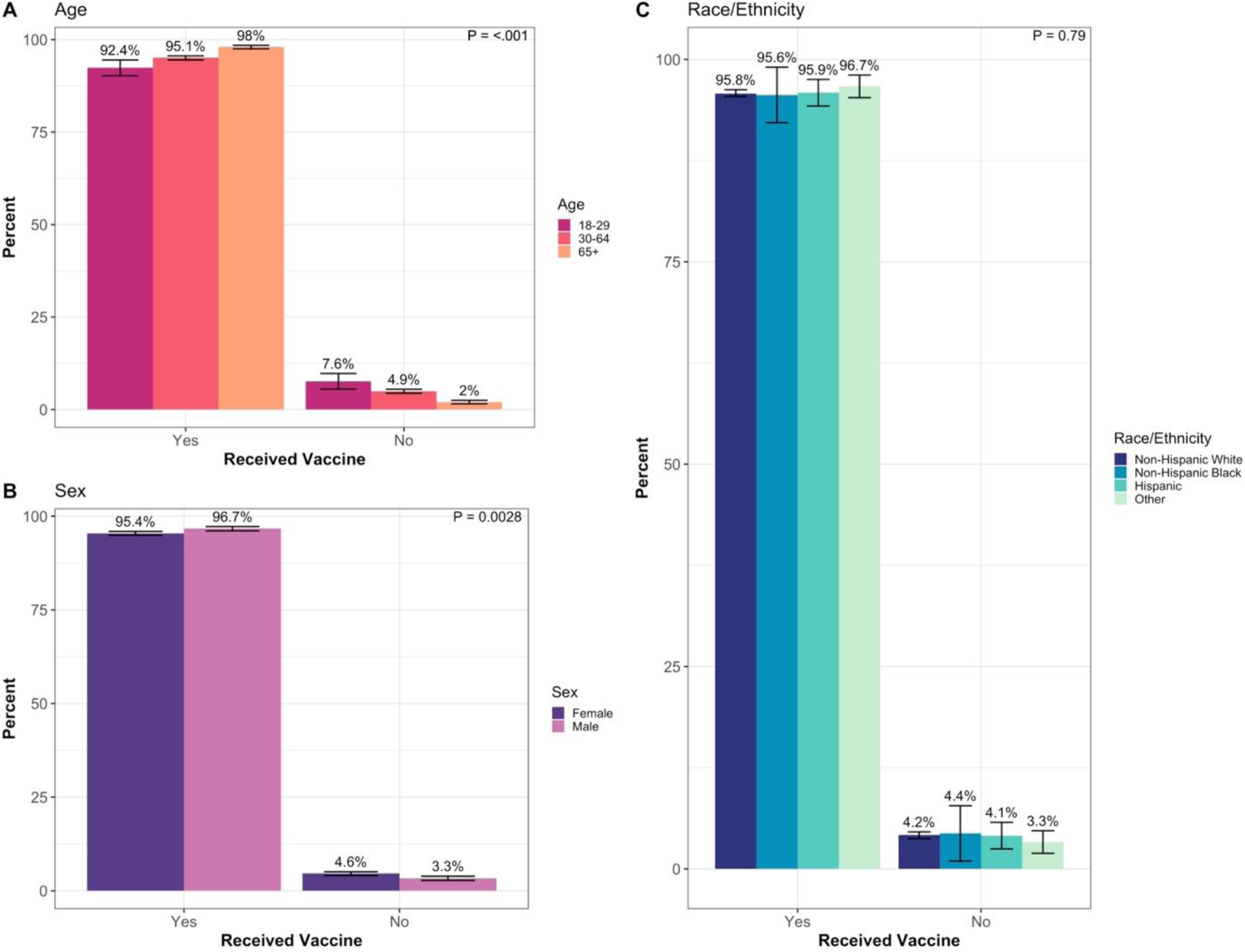
Vaccine uptake by A) age, B) sex, and C) race-ethnicity. P = *p*-value from Pearson’s chi-square test for different distributions across impact and demographic groups. Error bars indicate the 95% confidence interval for the percent point estimate.

### Demographics of COVID-19 Cases Captured by EHR versus Survey

We identified 11,472 COVID-19 positive cases from among 180,599 eligible Biobank participants: 1,366 from the survey (5.4% of respondents) and 10,639 from the EHR (5.9% of records); 533 cases were identified in both sources (Figure 6).

**Figure 6:**
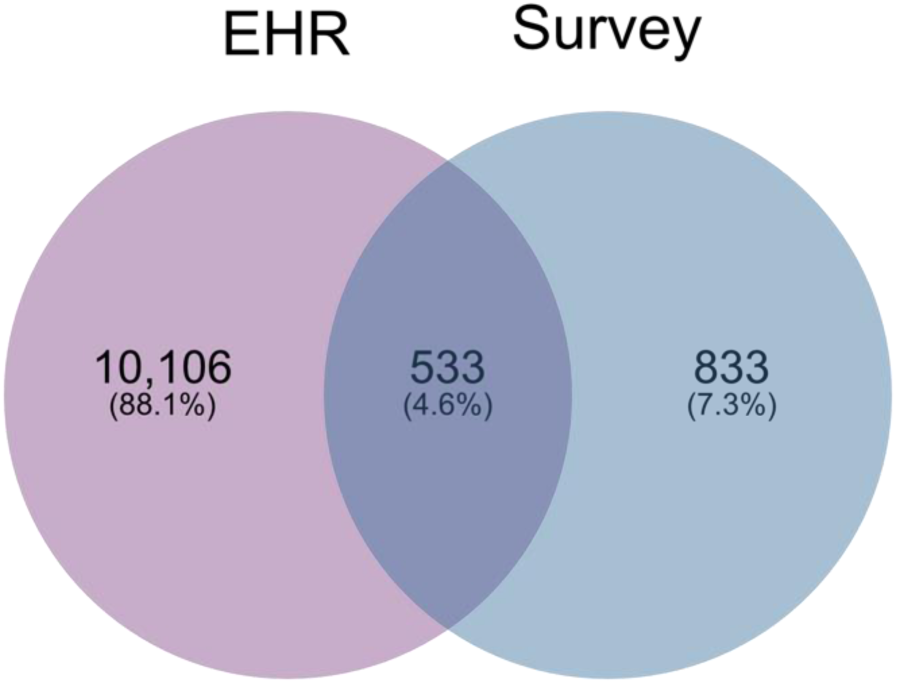
COVID-19 positive CCPM Biobank participants identified through the UCHealth EHR and the Survey.

In comparing COVID-19 cases from the EHR to those in the survey (**Figure 7**), we found that cases identified in the EHR were younger, with 17.2% of individuals in the 18-29 age group compared to 10.7% in the survey group (*P*<.001, Figure 7A). A higher percentage of cases identified in the EHR were Hispanic compared to survey cases (14.7% vs 9.2%, respectively, *P*<.001, Figure 7B). The EHR cases also had a slightly lower proportion of women (61.9%) compared to the survey group (66.0%); (*P* = 0.003, Figure 7C). The median income for the 3-digit zip code was the same, $69,900 in both groups. The median percent of the population who received a bachelor’s degree by 3-digit zip code was slightly lower in the EHR group (41.3%) compared to the survey (45.7%), *P*<.001.

**Figure 7:**
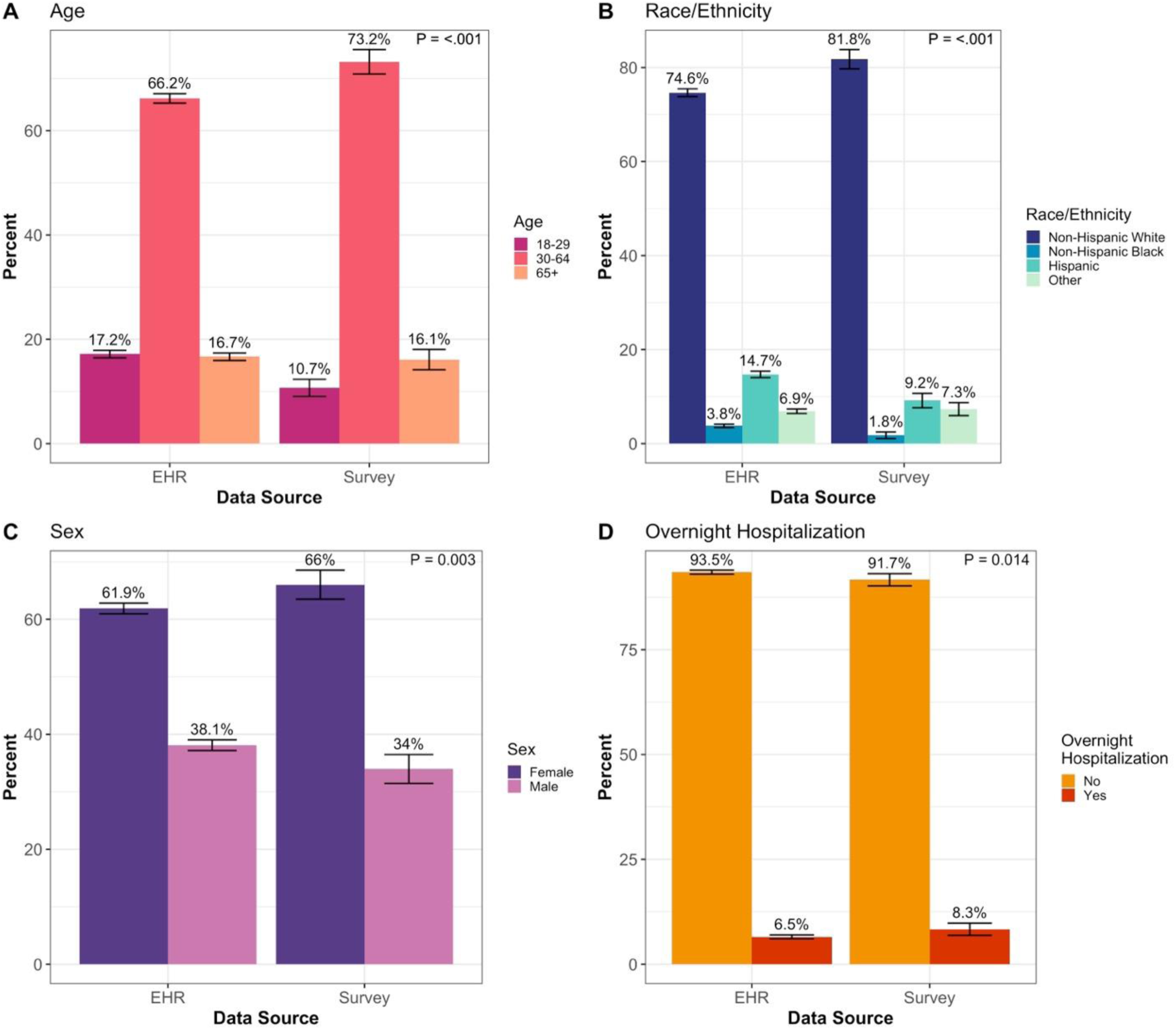
Comparison of COVID-19 cases captured in the EHR and the survey by A) age, B) race/ethnicity, C) sex, and D) COVID-19 related overnight hospitalization. P = *p*-value from Pearson’s chi-square test for different distributions across impact and demographic groups. Error bars indicate the 95% confidence interval for the percent point estimate.

### Comparison of COVID-19 Case Severity in EHR versus Survey

A higher percent of COVID-positive cases identified from the survey were hospitalized overnight (8.3%) compared to the EHR group (6.5%) (*P* = 0.01, Figure 7D). Using all-cause mortality data obtained from CDPHE vital statistics, 130 (2.3%) individuals in the EHR case group died, leading to a death rate of 1.2%. Four people in the survey case group died, with a death rate of 0.22%.

The EHR is a longitudinal data source, therefore we can capture COVID-19 cases on a continuing basis, whereas the survey reflects a point in time and can only identify individuals who had COVID-19 before they took the survey. Among 907 COVID-19 cases identified in the EHR who completed the survey but did not report a positive COVID-19 diagnosis in the survey, 41.8% reported receiving a negative COVID-19 test result (n = 379), and 58.2% had not taken a COVID-19 test and were presumed to be negative (n = 528). The majority of these individuals (80.7%) completed the survey before they were diagnosed with COVID-19 in the EHR (n = 732).

### COVID-19 Case and Hospitalization Discordance between the EHR and Survey

To quantify discordance of COVID-19 case status between the EHR and the survey we looked across our entire set of survey respondents (n = 25,063). We only counted a participant as “EHR COVID-19+” if the diagnosis was prior to taking the survey, not COVID-19 cases that happened after the survey was taken. While neither the survey nor the EHR are a gold standard for case classification, we can look at the discordance between them to identify the potential for misclassification. Overall, there were a total of 1,006 respondents discordant for COVID-19 case-status (4%). One hundred seventy-three individuals of the 25,063 individuals who took the survey were identified as COVID-19+ in the EHR but negative or not tested in the survey, leading to a discordance rate of 0.7% (**Table 2**). 833 individuals were identified as COVID-19+ in the survey but negative in the EHR, leading to a discordance rate of 3.3%.

**Table 2:** Case Status Misclassification Between the Survey and EHR.

|  | Survey COVID+ | Survey COVID Other<br>(Negative or Not Tested) | Total |
| --- | --- | --- | --- |
| EHR COVID+ | 533 | 173 | 706 |
| EHR COVID Other (Negative or Not Tested) | 833 | 23524 | 24357 |
| Total | 1366 | 23697 | 25063 |

To quantify discordance of hospitalization status in both the EHR and in the survey, we restricted it to individuals who responded to the survey and were either COVID-19 positive in the EHR or the survey (n = 2,273). EHR hospitalizations were only considered if they were prior to taking the survey. There were 6 individuals who were positive for hospitalization in the EHR but negative in the survey, a discordance rate of 0.3% (**Table 3**). There were 59 individuals who were positive for hospitalization in the survey who were negative in the EHR, a discordance rate of 2.6%.

**Table 3:** Hospitalization Misclassification Between the Survey and EHR.

|  | Survey Hospitalization + | Survey Hospitalization - | Total |
| --- | --- | --- | --- |
| EHR Hospitalization + | 49 | 6 | 55 |
| EHR Hospitalization - | 59 | 2159 | 2218 |
| Total | 108 | 2165 | 2273 |

## DISCUSSION

We found that the COVID-19 pandemic has had far-reaching and varying effects among our Biobank participants. Over 84% of the 25,063 survey respondents reported having one or more COVID-related symptoms since February 2020, 40% were tested for the virus, 13% of those tested were positive, and among positive cases, 45% sought medical care following their diagnosis. Our overall case positivity rate of 13% is comparable to those reported by other EHR-based retrospective studies conducted in 2020 and 2021 [8, 9]. However, our finding of higher positivity rates among our younger participants (aged 18-39 years; 20%) and Hispanics (19%) in our participants has not been reported previously and may reflect differences in reasons for testing in these groups (e.g. due to having symptoms or recent exposure vs. other reasons). Though not surprising that a large proportion of respondents reported having symptoms given the breadth of symptoms reported (e.g. runny nose, fever, body aches), it is notable that 40% of those with symptoms did not undergo testing nor seek medical care. It is likely that a percentage of this group had COVID-19 and would not be counted as such via public health surveillance efforts, which could lead to substantial underestimates of the true infection rate in the general population.

The vast majority of all survey respondents (75%) reported a negative impact from the COVID-19 pandemic—most commonly around mental health and family life. We found that females more often reported negative impacts than males in all domains–employment, family life, mental and physical health. This disproportionate negative impact on females is consistent with prior public health emergencies [10], including the 2016 Zika and 2014 Ebola outbreaks [11]. Among U.S. women, this has been described in several areas, including the healthcare workforce, reproductive health, drug development, gender-based violence, and mental health [12]. It is both notable and concerning that nearly 75% of younger adults (aged 18-29 years) reported negative impacts on their mental health, which was higher than for any other group. The younger end of this range captures members of Generation Z, who are more likely to report poor mental health compared to prior generations [13, 14]. However, they are also more likely to receive mental health therapy or treatment [13], and therefore may accept interventions to address the negative mental health consequences of the COVID-19 pandemic. Further, we found that negative impacts on employment were more commonly reported among Black participants. These findings highlight the breadth of negative impacts of this pandemic in our community, and reveal the disproportionate impact experienced by certain subgroups that should be targeted in future intervention efforts.

Our study population had a much higher vaccination rate compared to Colorado overall and the general US population. Over 95% of our survey participants are fully vaccinated compared to 76% of adults throughout Colorado [15]. Vaccination directly reduces likelihood of infection and severity of disease but it also has an indirect effect on society via reduced viral transmission and herd immunity. Because of this impact on others, getting vaccinated is considered a prosocial behavior [16-18]. Being a participant in a biobank has also been positively associated with prosocial behavior, as the individuals who participate in biobanks tend to be motivated by furthering research for the greater good [19, 20]. Since our study population only includes those who elected to be in the biobank and additionally, those who responded to the survey, these are likely individuals with high levels of prosocial behaviors, which likely explains the high vaccination rate. Given our highly self-selected study population, results may not generalize outside of the CCPM Biobank and UCHealth population. However, our ability to incorporate EHR data allows us to build a research population of biobank participants that is more representative of the entire patient population.

Differences between data captured in the EHR vs. those captured in the survey reveal the benefit of using both sources in combination. For example, mild cases with sub-clinical manifestations of infection that did not result in seeking care may be missing from the EHR but captured in a survey. The EHR is a longitudinal data source that collects clinical information on all patients diagnosed with and/or treated for COVID-19 within the UCHealth system irrespective of proclivity to participate in research or respond to surveys. As such, the EHR captured data from Biobank participants that the survey did not. Periodic analysis of EHR data will allow us to study COVID-19 reinfection and vaccine breakthrough cases over time. However, UCHealth is not a closed system and Biobank participants can receive care outside of UCHealth, so we recognize that not all individuals that were diagnosed or treated for COVID-19 are or will be captured in the EHR. We believe that the survey data more completely identifies individuals who did not seek healthcare or sought care outside of the UCHealth system, in particular patients who reported COVID-19 infection in the survey but had no corresponding record in the EHR.

COVID-19 has variable clinical presentations ranging from asymptomatic infections to severe symptoms that require hospitalization. We expected that COVID-19 patients identified in the EHR would be sicker on average than survey only cases, more likely to have severe COVID-19 and less likely to have asymptomatic infections [22, 23]. However, we found that there was a slightly higher percentage of COVID-19 hospitalizations among survey cases compared to EHR cases. This unexpected result may be explained, in part, by the fact that the survey likely undercounts deaths due to COVID-19, since the survey cannot identify anyone who died from COVID-19 unless they took the survey in between being diagnosed with COVID-19 and dying, whereas our clinical data warehouse (that combines EHR data with state vital statistics) will capture all deaths reported to CDPHE. With respect to participant demographics, it notable that a higher percentage of younger (18-29) and Hispanic/Latino positive cases were identified via the EHR vs. the survey. This may in part be explained by lower survey response rates in these groups. Hispanic/Latino individuals may have been less likely to take the survey because of language barriers (the survey was only in English), limited internet access or other structural barriers[24]. Lower participation among Hispanic individuals is consistent with observations in other outreach efforts [19806848] and is a limitation of the convenience survey design. Additionally, the Hispanic population in Colorado, as in many other states, had higher incidence of COVID-19 infections, hospitalizations, and death [4, 25-28], which may explain why they are more likely to be identified through the EHR.

A key strength to this study is our ability to leverage an existing, living resource in the CCPM Biobank and survey engine to assess the health and wellbeing of our participants in ways that are not highlighted by the EHR. Further, because participants consent to re-contact, we have the opportunity to follow up with sub-populations within our cohort to collect additional information and monitor outcomes such as re-infection and vaccine uptake. Although our overall response to the survey was sizeable, we acknowledge that the composition of the underlying patient population at UCHealth who enrolled in the Biobank, and differential response to the survey may have introduced some bias and limited the generalizability of our results.

## CONCLUSION

The combination of EHR and survey data provides a powerful opportunity to monitor and study the on-going effects of the COVID-19 pandemic in our communities. As the pandemic continues, there is a critical need for optimal COVID-19 case ascertainment in order to capture both mild and severe cases, and to monitor specific long-term outcomes such as post-acute sequelae of SARS-CoV-2 infection (PASC) or downstream breakthrough infections post-vaccination. In an open health system as is common in the United States, the development of a combined resource such as ours (with EHR and survey data) represents long-term potential for additional recruitment and follow-up as a critical complement to large-scale informatics-focused investigations such as the National COVID Cohort Collaborative[29]. As the pandemic continues, we anticipate that resources such as the CCPM Biobank and other biobanks will continue to be a key resource for ongoing data collection relevant to population health monitoring during the era of COVID-19 and other emerging public health issues.

## Data Availability

All data produced in the present study are available upon reasonable request to the authors

## ACKNOWLEDGMENTS

We would like to thank all participants in CCPM, as well as the entire CCPM team making these research initiatives a possibility in the pandemic. CCPM is supported by UCHealth and the University of Colorado Anschutz Medical Campus. We thank Eric Campbell for assistance on survey design. The research derivation of the EHR was made possible by the Health Data Compass Data Warehouse project (healthdatacompass.org). KMM and CRG are partially supported by R01HG011345. The REDCap database in this publication was supported by NIH/NCATS Colorado CTSA Grant Number UL1TR002535. The contents are the authors’ sole responsibility and do not necessarily represent official NIH views.

## ABBREVIATIONS

CCPM: Colorado Center for Personalized Medicine
EHR: Electronic Health Record

## REFERENCES

1. CDC. COVID-19. Centers for Disease Control and Prevention; [cited 2021 12/10/2021]; Available from: https://www.cdc.gov/coronavirus/2019-ncov/index.html.

2. Magesh S, John D, Li WT, Li Y, Mattingly-App A, Jain S, et al. Disparities in COVID-19 Outcomes by Race, Ethnicity, and Socioeconomic Status: A Systematic-Review and Meta-analysis. JAMA Netw Open. 2021 Nov 1;4(11):e2134147. PMID: 34762110. doi: 10.1001/jamanetworkopen.2021.34147.PMC8586903

3. Tenforde MW, Billig Rose E, Lindsell CJ, Shapiro NI, Files DC, Gibbs KW, et al. Characteristics of Adult Outpatients and Inpatients with COVID-19 - 11 Academic Medical Centers, United States, March-May 2020. MMWR Morb Mortal Wkly Rep. 2020 Jul 3;69(26):841-6. PMID: 32614810. doi: 10.15585/mmwr.mm6926e3.PMC7332092

4. Acosta AM, Garg S, Pham H, Whitaker M, Anglin O, O’Halloran A, et al. Racial and Ethnic Disparities in Rates of COVID-19-Associated Hospitalization, Intensive Care Unit Admission, and In-Hospital Death in the United States From March 2020 to February 2021. JAMA Netw Open. 2021 Oct 1;4(10):e2130479. PMID: 34673962. doi: 10.1001/jamanetworkopen.2021.30479.PMC8531997

5. Morlock R, Morlock A, Downen M, Shah SN. COVID-19 prevalence and predictors in United States adults during peak stay-at-home orders. PLoS One. 2021;16(1):e0245586. PMID: 33481900. doi: 10.1371/journal.pone.0245586.PMC7822538

6. International Common Disease Alliance. [January 10, 2022]; Available from: https://www.icda.bio/.

7. Harris PA, Taylor R, Thielke R, Payne J, Gonzalez N, Conde JG. Research electronic data capture (REDCap)--a metadata-driven methodology and workflow process for providing translational research informatics support. J Biomed Inform. 2009 Apr;42(2):377-81. PMID: 18929686. doi: 10.1016/j.jbi.2008.08.010.PMC2700030

8. Lindsay L, Secrest MH, Rizzo S, Keebler DS, Yang F, Tsai L. Factors associated with COVID-19 viral and antibody test positivity and assessment of test concordance: a retrospective cohort study using electronic health records from the USA. BMJ Open. 2021 Oct 1;11(10):e051707. PMID: 34598988. doi: 10.1136/bmjopen-2021-051707.PMC8488284

9. Sheehan MM, Reddy AJ, Rothberg MB. Reinfection Rates Among Patients Who Previously Tested Positive for Coronavirus Disease 2019: A Retrospective Cohort Study. Clinical Infectious Diseases. 2021;73(10):1882–6. doi: 10.1093/cid/ciab234

10. Smith J. Overcoming the ‘tyranny of the urgent’: integrating gender into disease outbreak preparedness and response. Gender & Development. 2019;27(2):355–69.

11. Davies SE, Bennett B. A gendered human rights analysis of Ebola and Zika: locating gender in global health emergencies. International Affairs. 2016;92(5):1041–60.

12. Connor J, Madhavan S, Mokashi M, Amanuel H, Johnson NR, Pace LE, et al. Health risks and outcomes that disproportionately affect women during the Covid-19 pandemic: A review. Soc Sci Med. 2020 Dec;266:113364. PMID: 32950924. doi: 10.1016/j.socscimed.2020.113364.PMC7487147

13. Association AP. 2018 Stress in America: Generation Z. Stress in America™ Survey. Washington, DC: American Psychological Association (APA), 2018.

14. Association AP. Stress in America™: One Year Later, A New Wave of Pandemic Health Concerns. Washington, DC: American Psychological Association (APA), 2021.

15. CDPHE. Colorado COVID-19 Vaccination Data. Colorado Department of Public Health and the Environment; [12/17/2021]; Available from: https://covid19.colorado.gov/vaccine-data-dashboard.

16. Böhm R, Betsch C, Korn L, Holtmann C. Exploring and Promoting Prosocial Vaccination: A Cross-Cultural Experiment on Vaccination of Health Care Personnel. Biomed Res Int. 2016;2016:6870984. PMID: 27725940. doi: 10.1155/2016/6870984.PMC5048021

17. Böhm R, Betsch C, Korn L. Selfish-rational non-vaccination: Experimental evidence from an interactive vaccination game. Journal of Economic Behavior & Organization. 2016 2016/11/01/;131:183–95. doi: https://doi.org/10.1016/j.jebo.2015.11.008

18. Cucciniello M, Pin P, Imre B, Porumbescu GA, Melegaro A. Altruism and vaccination intentions: Evidence from behavioral experiments. Soc Sci Med. 2021 Jul 13:114195. PMID: 34602309. doi: 10.1016/j.socscimed.2021.114195

19. Broekstra R, Aris-Meijer J, Maeckelberghe E, Stolk R, Otten S. Demographic and prosocial intrapersonal characteristics of biobank participants and refusers: the findings of a survey in the Netherlands. Eur J Hum Genet. 2021 Jan;29(1):11-9. PMID: 32737438. doi: 10.1038/s41431-020-0701-1.PMC7852517

20. Streicher SA, Sanderson SC, Jabs EW, Diefenbach M, Smirnoff M, Peter I, et al. Reasons for participating and genetic information needs among racially and ethnically diverse biobank participants: a focus group study. J Community Genet. 2011 Sep;2(3):153-63. PMID: 22109822. doi: 10.1007/s12687-011-0052-2.PMC3186034

21. CDPHE, Colorado State Emergency Operations Center, Testing & Notifications. [January 10, 2022]; Available from: https://covid19.colorado.gov/testing-notifications.

22. Violán C, Foguet-Boreu Q, Hermosilla-Pérez E, Valderas JM, Bolíbar B, Fàbregas-Escurriola M, et al. Comparison of the information provided by electronic health records data and a population health survey to estimate prevalence of selected health conditions and multimorbidity. BMC Public Health. 2013 Mar 21;13:251. PMID: 23517342. doi: 10.1186/1471-2458-13-251.PMC3659017

23. Mizrahi B, Shilo S, Rossman H, Kalkstein N, Marcus K, Barer Y, et al. Longitudinal symptom dynamics of COVID-19 infection. Nat Commun. 2020 Dec 4;11(1):6208. PMID: 33277494. doi: 10.1038/s41467-020-20053-y.PMC7718370

24. Jang M, Vorderstrasse A. Socioeconomic Status and Racial or Ethnic Differences in Participation: Web-Based Survey. JMIR Res Protoc. 2019 Apr 10;8(4):e11865. PMID: 30969173. doi: 10.2196/11865.PMC6479282

25. Podewils LJ, Burket TL, Mettenbrink C, Steiner A, Seidel A, Scott K, et al. Disproportionate Incidence of COVID-19 Infection, Hospitalizations, and Deaths Among Persons Identifying as Hispanic or Latino - Denver, Colorado March-October 2020. MMWR Morb Mortal Wkly Rep. 2020 Dec 4;69(48):1812-6. PMID:33270613. doi: 10.15585/mmwr.mm6948a3.PMC7714035

26. Moore JT, Ricaldi JN, Rose CE, Fuld J, Parise M, Kang GJ, et al. Disparities in Incidence of COVID-19 Among Underrepresented Racial/Ethnic Groups in Counties Identified as Hotspots During June 5-18, 2020 - 22 States, February-June 2020. MMWR Morb Mortal Wkly Rep. 2020 Aug 21;69(33):1122-6. PMID: 32817602. doi: 10.15585/mmwr.mm6933e1.PMC7439982

27. Romano SD, Blackstock AJ, Taylor EV, El Burai Felix S, Adjei S, Singleton CM, et al. Trends in Racial and Ethnic Disparities in COVID-19 Hospitalizations, by Region - United States, March-December 2020. MMWR Morb Mortal Wkly Rep. 2021 Apr 16;70(15):560-5. PMID: 33857068. doi: 10.15585/mmwr.mm7015e2.PMC8344991

28. Mackey K, Ayers CK, Kondo KK, Saha S, Advani SM, Young S, et al. Racial and Ethnic Disparities in COVID-19-Related Infections, Hospitalizations, and Deaths : A Systematic Review. Ann Intern Med. 2021 Mar;174(3):362-73. PMID: 33253040. doi: 10.7326/m20-6306.PMC7772883

29. National COVID Cohort Collaborative (N3C). [January 10, 2022]; Available from: https://ncats.nih.gov/n3c.

