## Supplemental Materials for "COVID-19 Surveillance in the Biobank at the Colorado Center for Personalized Medicine"

**Multimedia Appendix 1:** Characteristics of Participants in the Biobank at the Colorado Center for Personalized Medicine Compared to the UC Health System

|  | **Biobank Participants*** | **UC Health**** |
| --- | --- | --- |
| **Characteristics** | **N=180,599** | **N=2,669,633** |
| **Sex** |  |  |
| Female | 60% | 54% |
| Male | 41% | 46% |
| Unknown | 0% | 0% |
| **Ethnicity** |  |  |
| Not Hispanic or Latino | 88% | 72% |
| Unknown | 3% | 15% |
| Hispanic or Latino | 9% | 13% |
| **Race** |  |  |
| White | 83% | 70% |
| Unknown | 10% | 22% |
| Black or African American | 4% | 6% |
| Asian | 2% | 2% |
| American Indian or Alaska Native | 0% | 0% |
| Native Hawaiian or Other Pacific Islander | 0% | 0% |
| **Age** |  |  |
| 18-29 | 13% | 19% |
| 30-39 | 22% | 19% |
| 40-49 | 17% | 16% |
| 50-59 | 16% | 15% |
| 60-69 | 17% | 15% |
| 70-79 | 12% | 10% |
| 80+ | 3% | 7% |
| Unknown | 0% | - |
| **Biobank at the CCPM enrollment as of 05/31/2021* | | |
| ***UC Health patient population (>= 18 years of age) accessed from TriNetX, Data updated 6/24/2021* | | |

**Multimedia Appendix 2:** COVID-19 Survey Instrument Administered in 2020
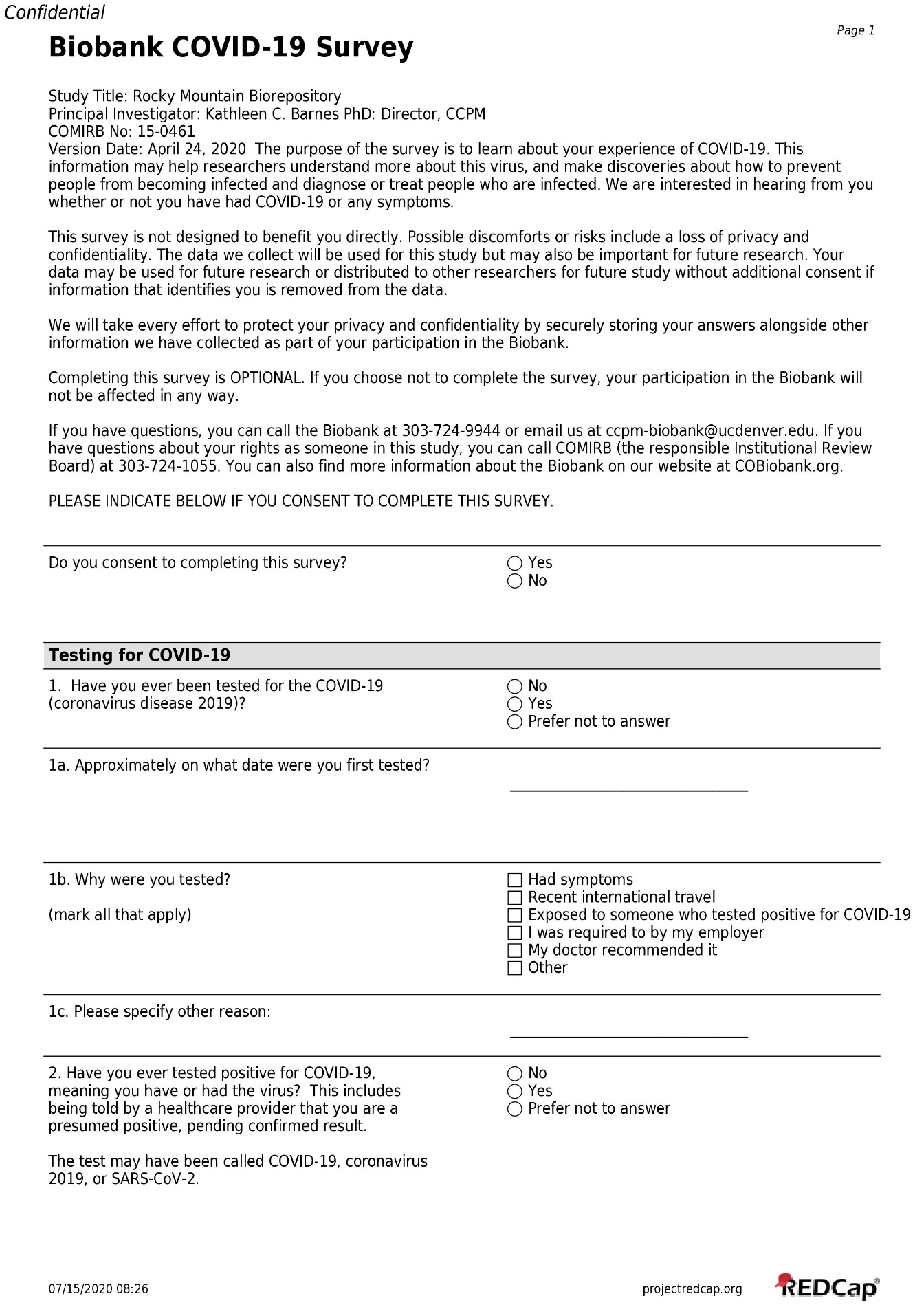

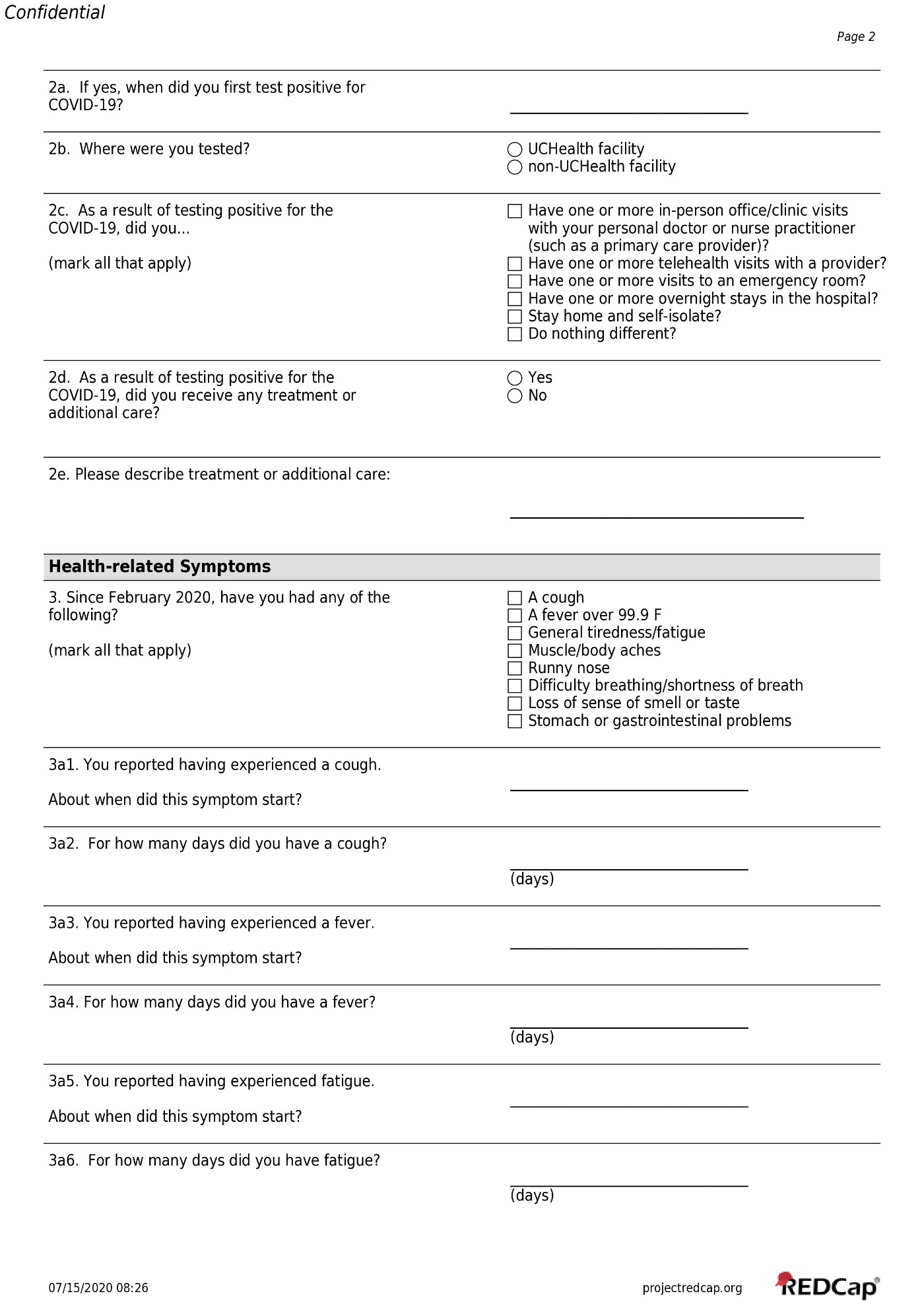

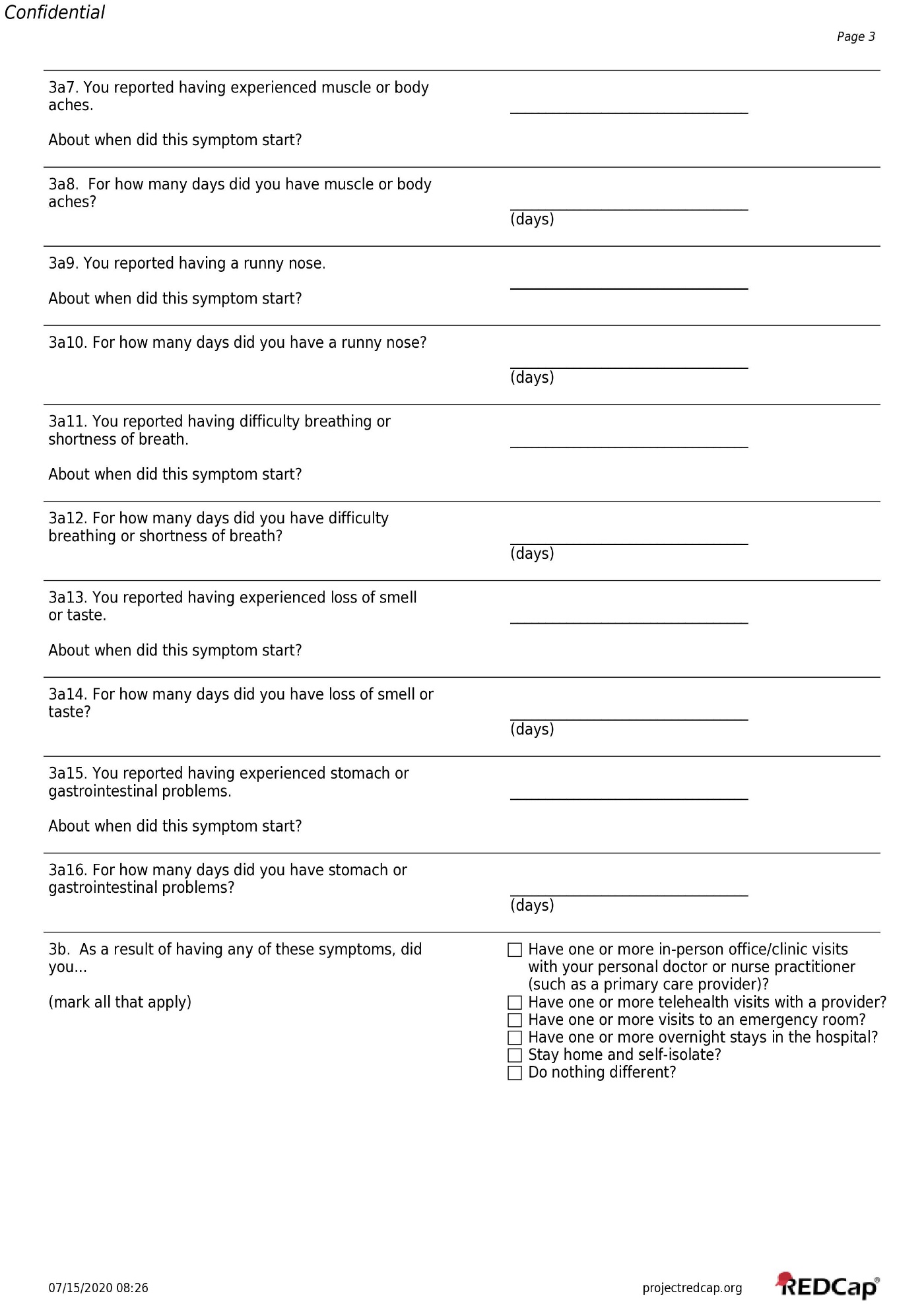

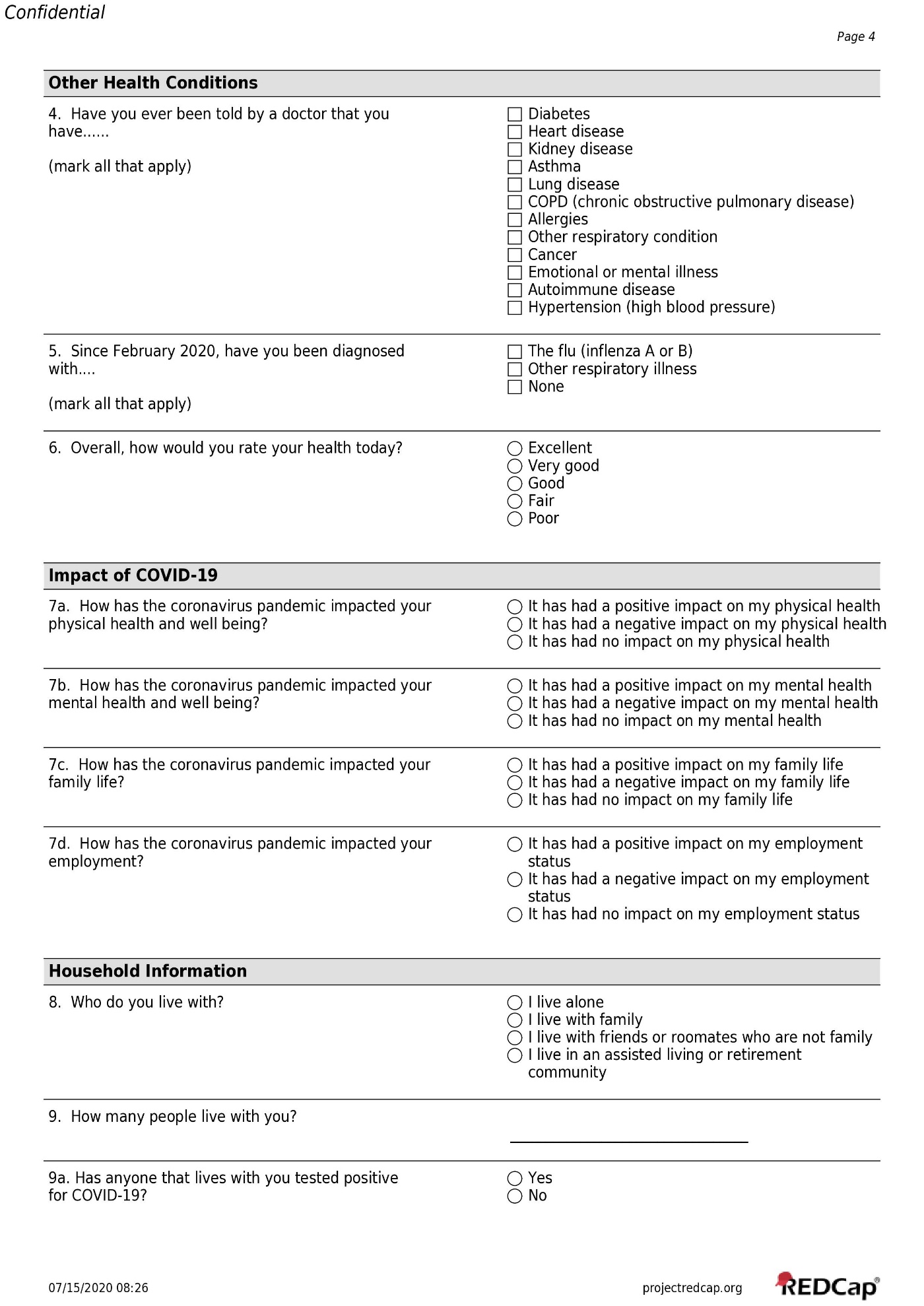

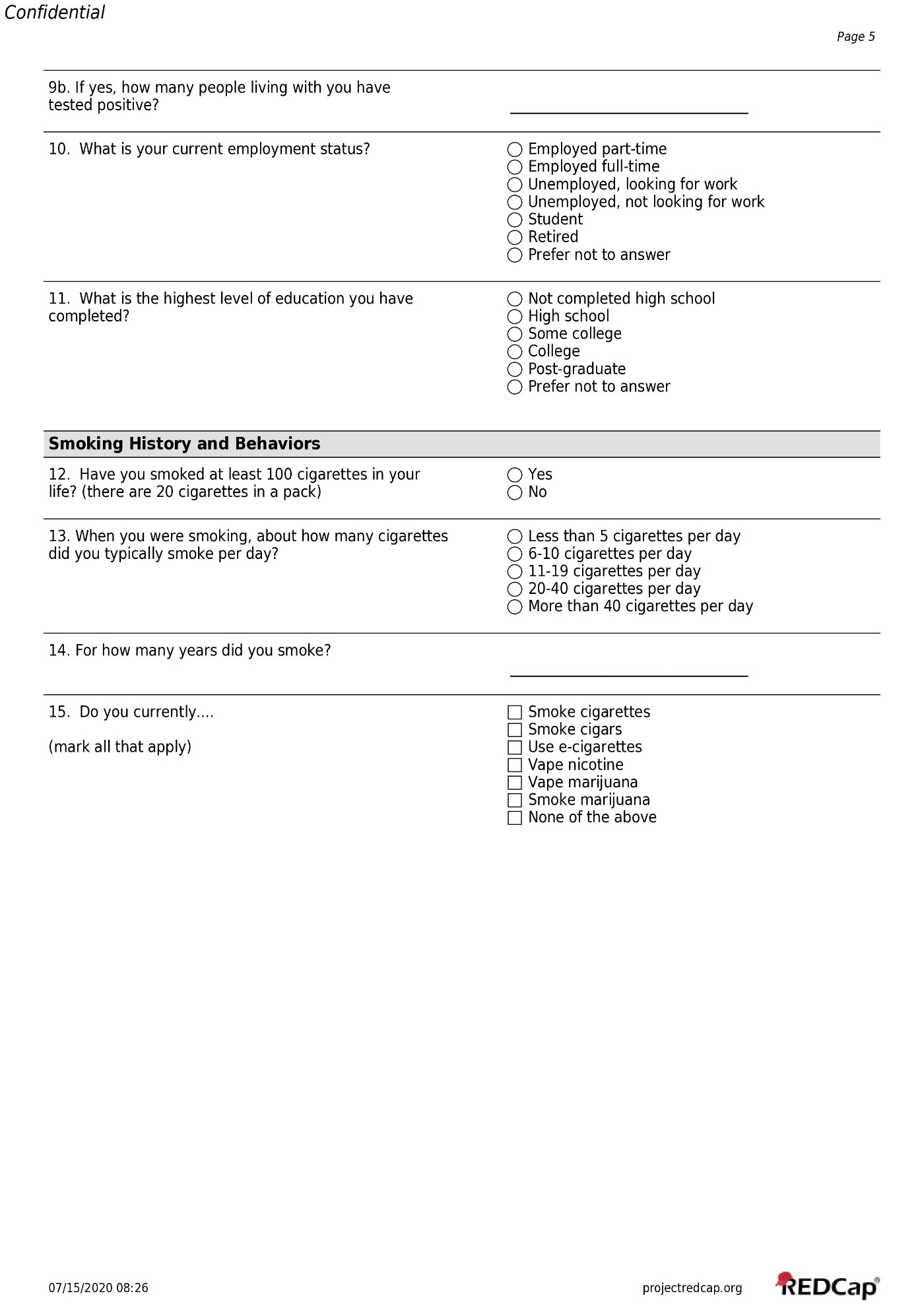

**Multimedia Appendix 3:** COVID-19 Survey Instrument Administered in 2021
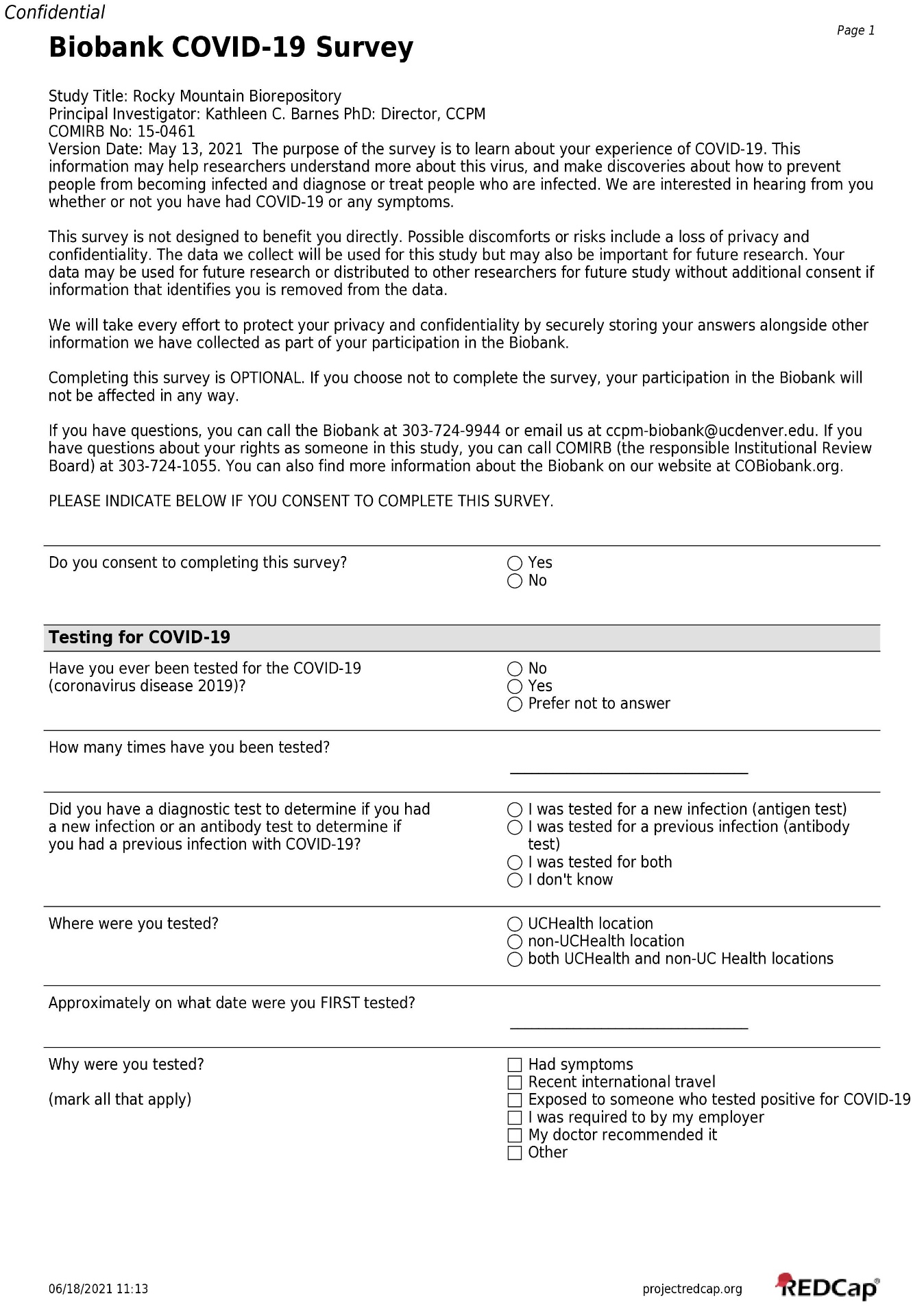

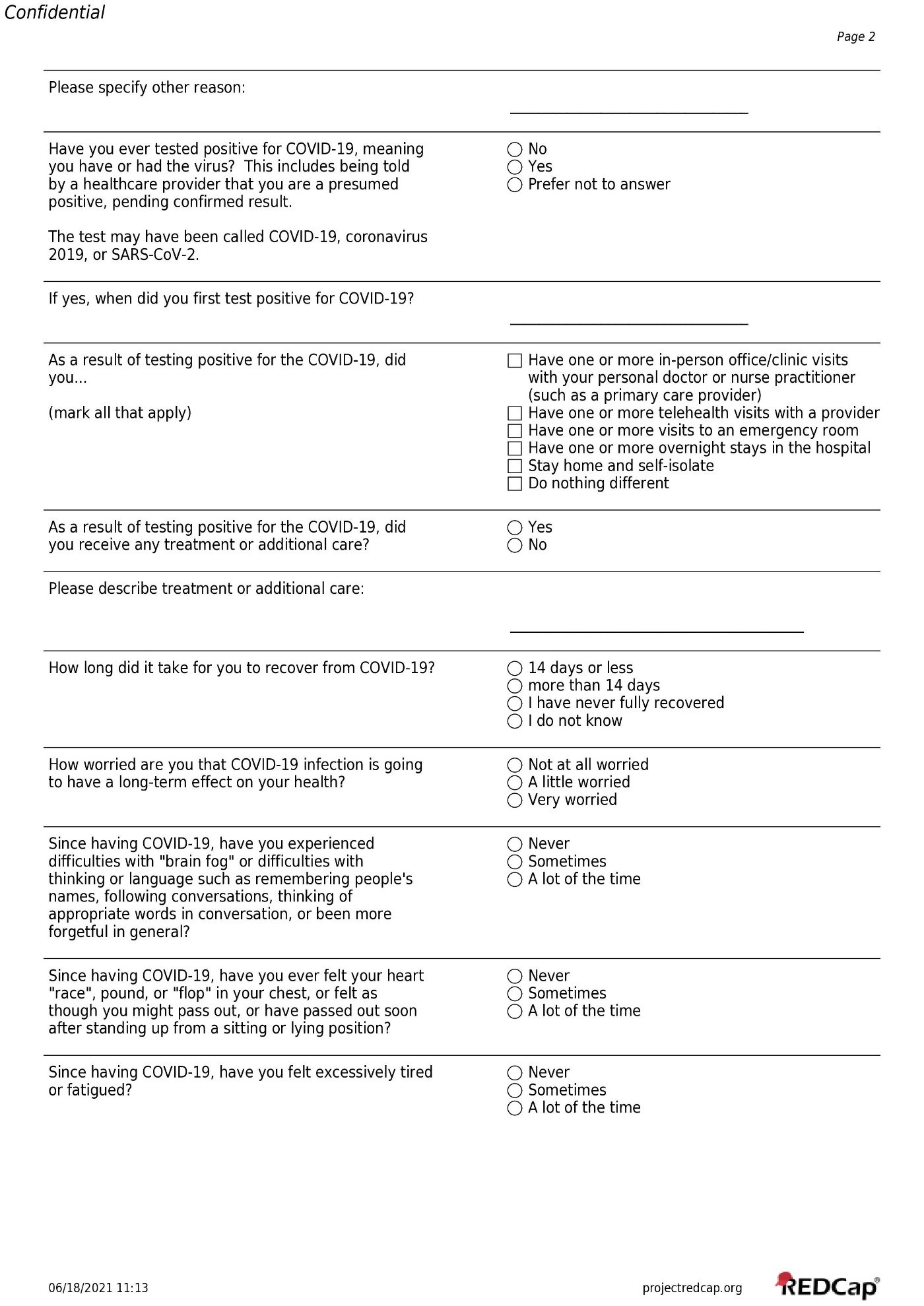

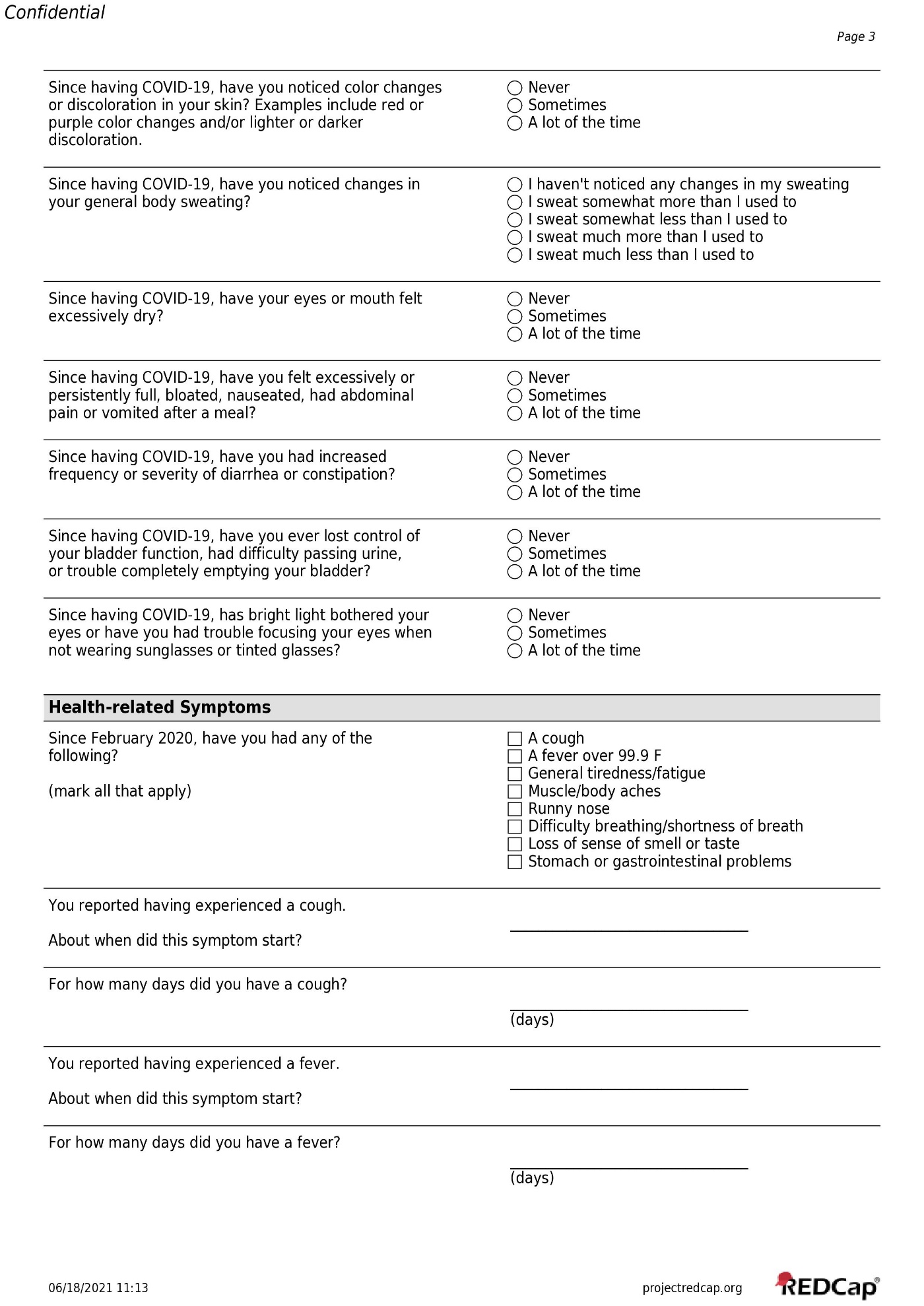

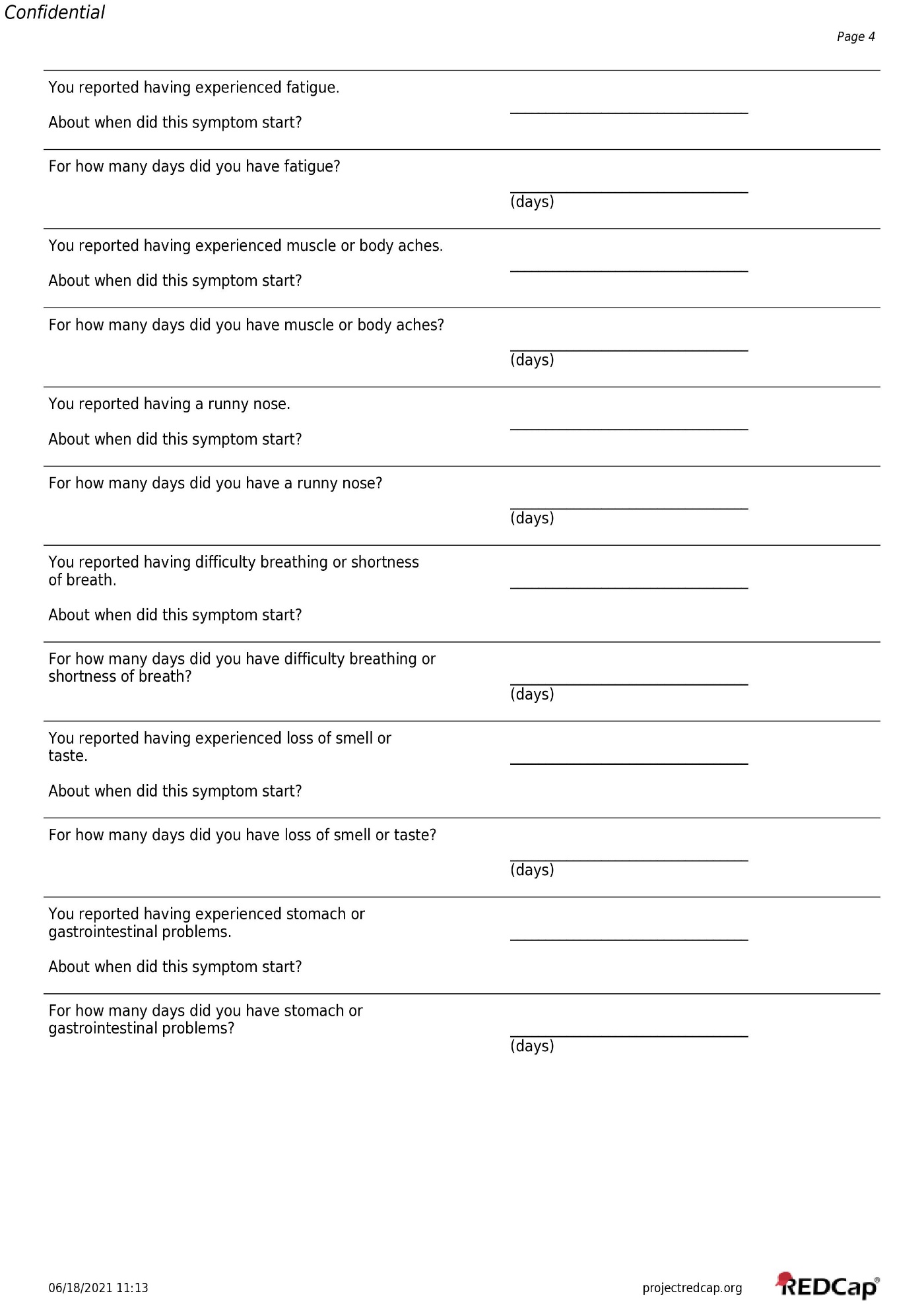

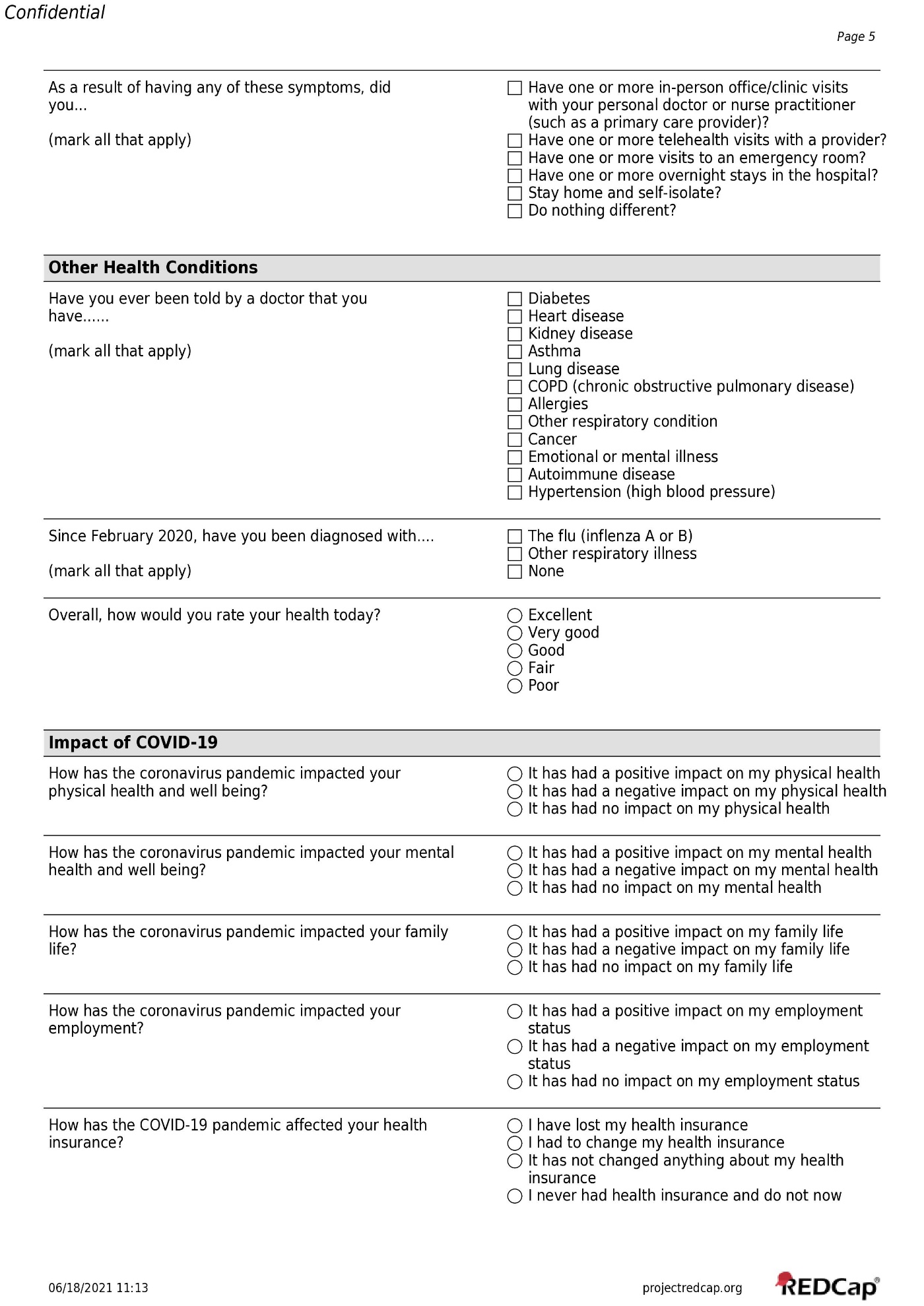

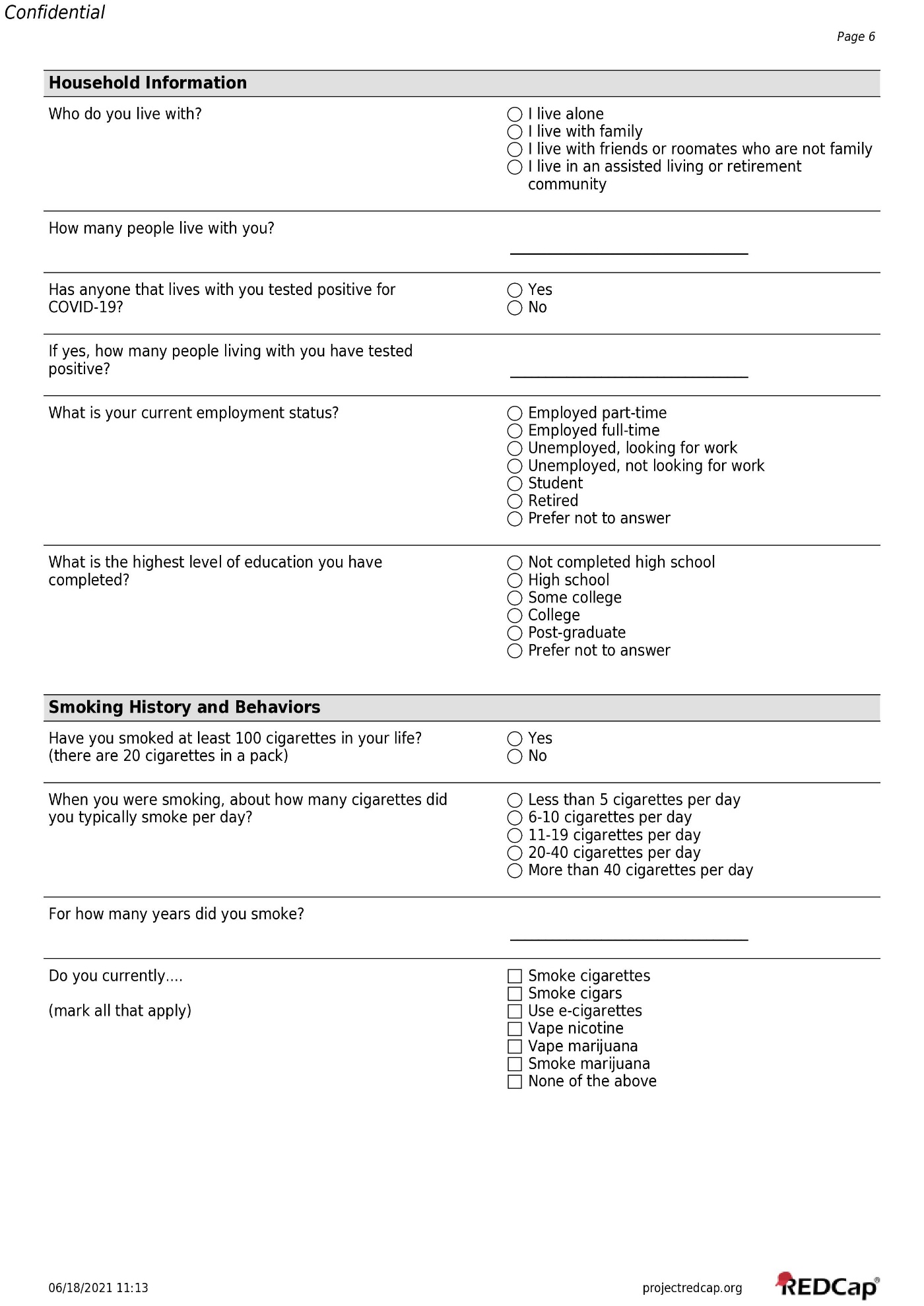

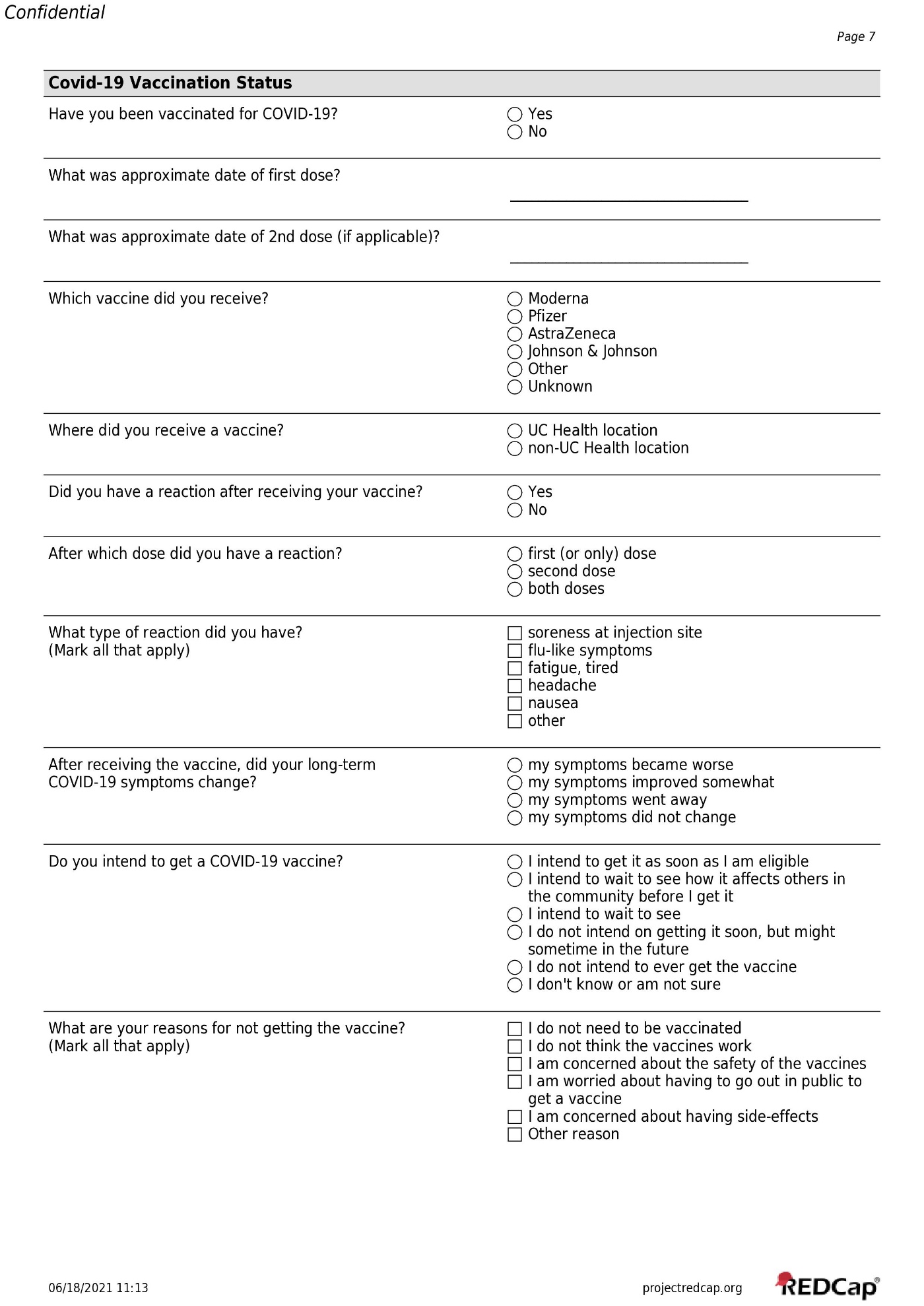

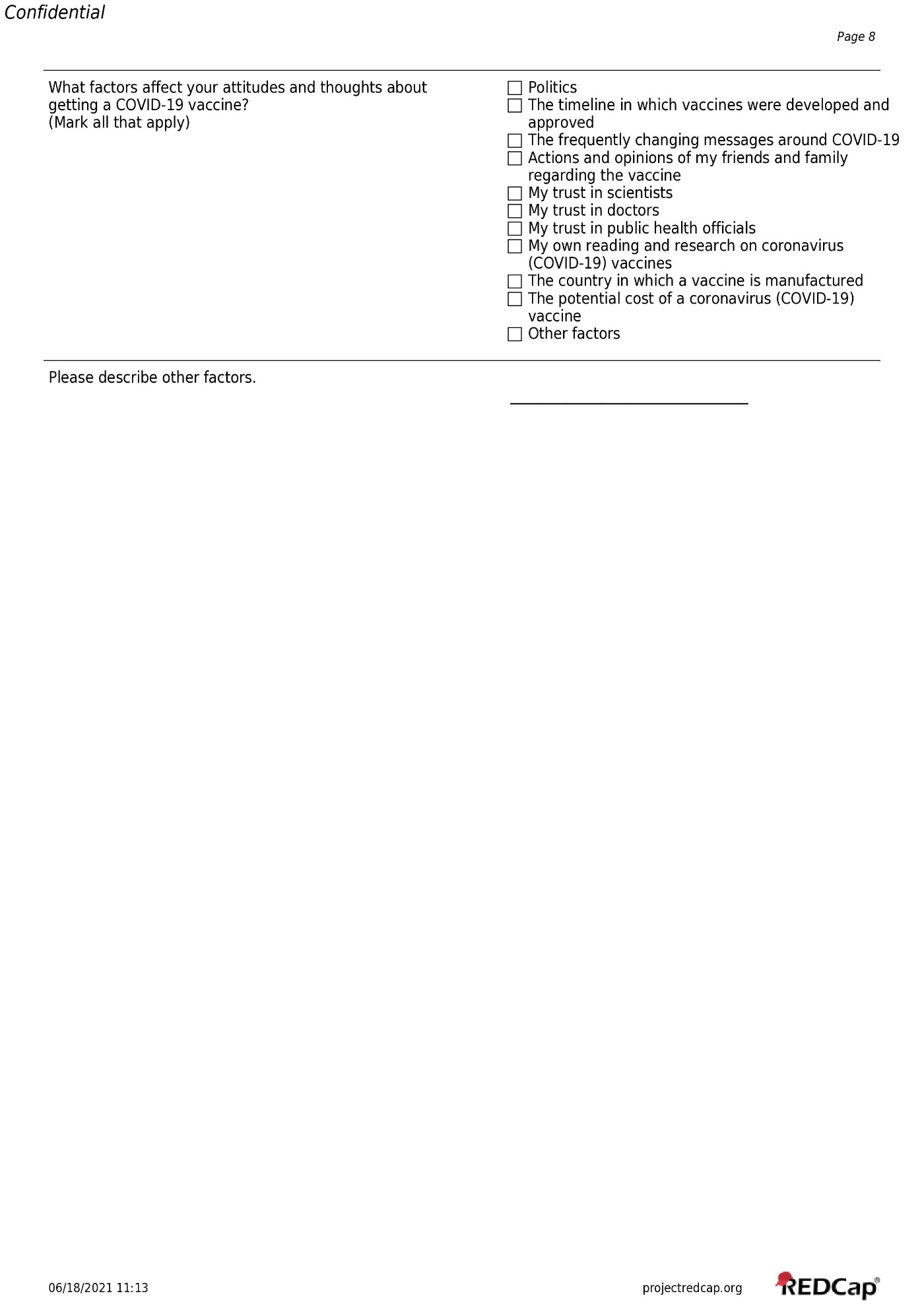

**Multimedia Appendix 4:** COVID-19 specific encounter primary diagnoses used in addition to U07.1 to identify “EHR-confirmed case” from UC Health EHR data.

"2019 novel coronavirus disease (COVID-19)",

"Acute respiratory disease due to COVID-19 virus",

"Acute respiratory distress syndrome (ARDS) due to COVID-19 virus",

"COVID-19 virus detected",

"Encephalopathy due to COVID-19 virus",

"Gastroenteritis due to COVID-19 virus",

"Myocarditis due to COVID-19 virus",

"Otitis media due to COVID-19 virus",

"Pneumonia due to COVID-19 virus",

"Real time reverse transcriptase PCR positive for COVID-19 virus",

"Upper respiratory tract infection due to COVID-19 virus"

**Multimedia Appendix 5:** Specific encounter primary diagnoses used in conjunction with hospitalization, timing of hospitalization, and COVID-19 case definitions to identify “EHR-hospitalization” from UC Health EHR data.

"COVID-19",

"Other viral diseases complicating childbirth",

"Viral infection, unspecified",

"Pneumonia due to other specified infectious organisms",

"Sepsis due to COVID-19 (HC code)",

"Other viral pneumonia",

"Nausea and vomiting",

"Cerebral infarction due to unspecified occlusion or stenosis of right posterior cerebral artery",

"Acute on chronic systolic (congestive) heart failure",

"Acute on chronic combined systolic (congestive) and diastolic (congestive) heart failure",

"Other transient cerebral ischemic attacks and related syndromes",

"Acute kidney failure with tubular necrosis",

"Acute on chronic respiratory failure, unspecified whether with hypoxia or hypercapnia (HC code)",

"Acute embolism and thrombosis of left femoral vein",

"AMS (altered mental status)",

"Other coronavirus as the cause of diseases classified elsewhere",

"Single subsegmental pulmonary embolism without acute cor pulmonale",

"Other specified sepsis",

"Other viral diseases complicating pregnancy, third trimester",

"Other pulmonary embolism without acute cor pulmonale",

"Acute respiratory failure with hypoxia",

"Other encephalopathy",

"Other diseases of pharynx",

"Heart failure, unspecified",

"Acute embolism and thrombosis of right iliac vein",

"Acute pulmonary embolism without acute cor pulmonale, unspecified pulmonary embolism type (HC code)",

"Saddle embolus of pulmonary artery with acute cor pulmonale",

"Contact with and (suspected) exposure to other viral communicable diseases",

"Acute combined systolic (congestive) and diastolic (congestive) heart failure",

"Hypoxemia",

"Acute on chronic diastolic (congestive) heart failure",

"Shortness of breath",

"Other pulmonary embolism with acute cor pulmonale",

"Pneumonia, unspecified organism",

"Suspected COVID-19 virus infection",

"Acute respiratory distress syndrome",

"Acute upper respiratory infection, unspecified",

"Sepsis (HC code)",

"ST elevation (STEMI) myocardial infarction involving left circumflex coronary artery",

"Non-ST elevation (NSTEMI) myocardial infarction",

"COVID-19 virus infection",

"Cerebral infarction due to embolism of other cerebral artery",

"Pneumonia due to COVID-19 virus",

"Severe sepsis with septic shock (CODE) (HC code)",

"Embolism and thrombosis of iliac artery",

"Acute and chronic respiratory failure with hypoxia",

"Hypoxia",

"Cerebral infarction due to unspecified occlusion or stenosis of left carotid arteries",

"ST elevation (STEMI) myocardial infarction involving other coronary artery of inferior wall",

"Acute respiratory failure with hypoxia (HC code)",

"Cerebral infarction due to embolism of right middle cerebral artery",

"ST elevation (STEMI) myocardial infarction involving other coronary artery of anterior wall",

"Other specified respiratory disorders",

"Other cerebral infarction",

"Unspecified acute lower respiratory infection",

"Other chest pain",

"Viral pneumonia, unspecified",

"Other viral infections of unspecified site",

"Sepsis, unspecified organism",

"Other viral diseases complicating pregnancy, first trimester",

"Multiple subsegmental pulmonary emboli without acute cor pulmonale",

"Cerebral infarction, unspecified",

"Transient cerebral ischemic attack, unspecified",

"Diseases of the respiratory system complicating childbirth",

"Cerebral infarction due to unspecified occlusion or stenosis of unspecified carotid artery"

**Multimedia Appendix 6:** Characteristics of Biobank participants by COVID-19 survey response.

|  | **Total Biobank*** | **Response** | **Non-Response** | ***P-value*** |
| --- | --- | --- | --- | --- |
| **Characteristics** | **N=180,599** | **N=25,075** | **N=155,524** |  |
| **Age, mean (SD)** | 49.5 (17.0) | 55.0 (15.8) | 48.6 (17.0) | <.001 |
| **Age, n (%)** |  |  |  | <.001 |
| 18-29 | 23790 (13.2%) | 1504 (6.0%) | 22286 (14.3%) |  |
| 30-39 | 38825 (21.5%) | 3893 (15.5%) | 34932 (22.5%) |  |
| 40-49 | 30872 (17.1%) | 3824 (15.3%) | 27048 (17.4%) |  |
| 50-59 | 29248 (16.2%) | 4472 (17.8%) | 24776 (15.9%) |  |
| 60-69 | 30722 (17.0%) | 6180 (24.6%) | 24542 (15.8%) |  |
| 70-79 | 21025 (11.6%) | 4357 (17.4%) | 16668 (10.7%) |  |
| 80+ | 5991 (3.3%) | 836 (3.3%) | 5155 (3.3%) |  |
| Missing | 126 (0.1%) | 9 (0.0%) | 117 (0.1%) |  |
| **Sex, n (%)** |  |  |  | <.001 |
| Female | 107402 (59.5%) | 15695 (62.6%) | 91707 (59.0%) |  |
| Male | 73061 (40.5%) | 9368 (37.4%) | 63693 (41.0%) |  |
| Unknown | 136 (0.1%) | 12 (0.0%) | 124 (0.1%) |  |
| **Race-Ethnicity, n (%)** |  |  |  | <.001 |
| Non-Hispanic White | 141765 (78.5%) | 21917 (87.4%) | 119848 (77.1%) |  |
| Non-Hispanic Black | 6816 (3.8%) | 308 (1.2%) | 6508 (4.2%) |  |
| Hispanic | 16283 (9.0%) | 1272 (5.1%) | 15011 (9.7%) |  |
| Asian | 3628 (2.0%) | 329 (1.3%) | 3299 (2.1%) |  |
| American Indian and Alaska Native | 513 (0.3%) | 47.0 (0.2%) | 466 (0.3%) |  |
| Native Hawaiian and Other Pacific Islander | 288 (0.2%) | 15 (0.1%) | 273 (0.2%) |  |
| Non-Hispanic Other | 11180 (6.2%) | 1178 (4.7%) | 10002 (6.4%) |  |
| Unknown | 126 (0.1%) | 9 (0.0%) | 117 (0.1%) |  |
| **Genetic Data** |  |  |  | <.001 |
| N | 147647 (81.8%) | 20194 (80.5%) | 127453 (82.0%) |  |
| Y | 32952 (18.2%) | 4881 (19.5%) | 28071 (18.0%) |  |
| **Medican Income**, mean (SD)** | 75000 (24000) | 76700 (23800) | 74700 (24000) | <.001 |
| **Percent Above Bachelors Education**, mean (SD)** | 43.7 (17.4) | 46.6 (16.6) | 43.3 (17.5) | <.001 |
| **Biobank participants as of May 31, 2021, Responses to survey received by 10/31/2021* | | | | |
| ***Based on 3-digit zip code of residence* | | | | |
